# The Association Between Cognitive Ability and Body Mass Index: A Sibling-Comparison Analysis in Four Longitudinal Studies

**DOI:** 10.1101/2022.08.05.22277792

**Authors:** Liam Wright, Neil M Davies, David Bann

## Abstract

**Background:** Body mass index (BMI) and obesity rates have increased sharply since the 1980s. While multiple epidemiologic studies have found higher adolescent cognitive ability is associated with lower adult BMI, residual and unobserved confounding due to family background may explain these associations. We used a sibling design to test this association accounting for confounding factors shared within households.

**Methods:** We used data from four cohort studies: the National Longitudinal Study of Youth 1979 (NLSY-79), the NLSY-79 Children and Young Adult, the NLSY 1997 (NLSY-97) and the Wisconsin Longitudinal Study (WLS); a total of 12,250 siblings from 5,602 households. We used random effects within-between (REWB) and residualized quantile regression (RQR) models to compare between- and within-family estimates of the association between adolescent cognitive ability and adult BMI (20-64 years).

**Results:** In REWB models, moving from 0th to 100th percentile of adolescent cognitive ability was associated with −1.89 kg/m^2^ (95% CI = −2.41, −1.37) lower BMI between families. Adjusting for family socioeconomic position reduced the association to −1.23 (−1.79, −0.66) points. However, within families the association was just −0.13 (−0.70, 0.45) points. This pattern of results was found across multiple specifications, including analyses conducted in separate cohorts, models examining age-differences in association, and in RQR models examining the association across the distribution of BMI.

**Conclusion:** The association between high adolescent cognitive ability and low adult BMI was substantially smaller in within-family compared with between-family analysis. The well-replicated associations between cognitive ability and subsequent BMI may largely reflect confounding by family background factors.

## Introduction

Obesity rates have increased dramatically across developed countries since the 1980s [1], with significant consequences for public health and national economies [2]. Explanations for rising obesity rates have highlighted the ‘Big-Two’, changes to diets – resulting from wider availability of high energy density foods – and reductions in physical activity, though several other factors have also been proposed [3,4]. Despite numerous policy efforts to tackle rising obesity rates [5,6], and increasing public knowledge of its deleterious consequences, obesity remains a leading cause of disability and mortality [6].

While the prevalence of obesity (body mass index [BMI] ≥ 30 kg/m^2^) has increased, changes across the full distribution of BMI have not been as pronounced, with relatively little change in the prevalence of underweight and small increases in median BMI [7–11]. This suggests that individuals differ in their exposure to the causes of obesity or their susceptibility to these causes [12]; estimates of the heritability of BMI from twin studies range 47%-90% [13]. Thus, while obesity has increased in response to societal change, individual factors still have a role. Identifying these factors is paramount if the causes of the obesity epidemic to be understood and potential means to address it are to be to be found.

One characteristic that has been proposed to influence BMI – and health more generally – is cognitive ability. The field of cognitive epidemiology [14] has gathered substantial evidence that higher cognitive ability is associated with better health outcomes, including lower rates of mortality and major chronic diseases [15]. Cognitive ability is thought to affect health for two main reasons [15–17]. First, cognitive ability is related to higher socioeconomic position (SEP) – for instance, greater earnings and higher educational attainment [18] – and cognition may operate indirectly through this factor. In the present setting, by earning larger incomes, individuals with higher cognitive ability are likely to have greater access to healthier and more varied diets and to live in safer, more walkable neighbourhoods. Second, cognitive ability is argued to operate more directly by increasing individuals’ ability to understand and use nutritional, and other health-relevant, information. Correctly using nutritional labelling, for example, requires integrating existing nutritional knowledge with literacy and numeracy skills to decode information and make inferences about the healthiness of food items [19]. Health literacy was described in a 2022 WHO report as an “unrecognized obesity determinant” [6] and many people struggle to understand food labels and other health information [20–22]. Importantly, there is considerable overlap between the concepts of literacy and general cognitive ability [16]. Measures of health literacy may have little incremental validity over measures of intelligence [23].

Many studies have examined the association between cognitive ability and BMI [24]. Some have adopted a longitudinal design and investigated the link between childhood or adolescent cognitive ability and adult BMI, given the possibility of reverse causality in cross-sectional data [25]. Existing longitudinal studies span multiple countries, including several Scandinavian countries marked by low social inequality. Most [26–39] but not all [39] find that individuals with higher cognitive ability have lower BMI and obesity rates in adulthood. Effect sizes are typically stronger for obesity than (mean) BMI [31,34,38,39] and there is evidence that associations are stronger at older ages [29,31,35]. The results from longitudinal studies are consistent with Mendelian Randomization (MR) evidence from samples of unrelated individuals showing an association between genetic predisposition to high (adult) cognitive ability and lower BMI [40].

Though associations between cognitive ability and BMI are widely found, results in observational studies could be explained by unobserved or residual confounding. Specifically, associations may be driven by differences in early family environments, such as early SEP and parenting practices, that are either unmeasured or difficult to measure with available data (see Figure S1 for a directed acyclic graph): childhood and adolescent BMI (which are correlated with adult BMI [41]) are related to maternal cognitive ability [42] and there are strong socioeconomic gradients in BMI and obesity [43,44]. *Dynastic* effects operating via parental genetics to offspring BMI could partly explain genetic associations, too [40]. Existing studies on the link between cognitive ability and BMI have attempted to capture early SEP by controlling for one or a few high-level variables (e.g., parent’s years of education, occupational prestige, and self-reported income), but these have had limited granularity and spanned a narrow age range of childhood or adolescence [26,30,31,45].

Sibling comparison designs offer one approach to mitigate such confounding. On the assumption that early family environments are shared between siblings, comparing outcomes within sibships removes bias arising from family background (including shared dynastic effects). To our knowledge, no studies have used a sibling design to examine associations between early life cognition and BMI during adulthood. In this study, we combined sibling data from four cohort studies to examine the within-family association between adolescent cognition and adult BMI; large samples are required to achieve adequate statistical power in sibling designs [46]. Given existing evidence that associations are stronger for obesity than BMI, we used a novel statistical approach – Residualized Quantile Regression [47] – to examine (within-family) associations with cognitive ability across the BMI distribution, rather than just the (conditional) mean.

## Methods

### Participants

We used data from four cohort studies, each following participants from adolescence across adulthood and containing measures of adolescent cognitive ability and adult BMI: the National Longitudinal Surveys of Youth, 1979 and 1997 (NLSY-79 and NLSY-97, respectively), the NLSY-79 Child and Young Adults (NLSY-79 C/YA), and the Wisconsin Longitudinal Study (WLS). These cohorts are described in detail in the Supplementary Information. Briefly, the NLSY-79 [48] and NLSY-97 [49] are ongoing studies of young people in the United States that began in 1979 and 1997, respectively. Recruitment to the studies took place at the household level meaning multiple sibling sets are included. Beside a nationally representative sample, the NLSY-79 and NLSY-97 also include an oversample of ethnic minority (and, for the NLSY-79, economically disadvantaged) individuals, which we treated as separate cohorts. The NLSY-79 C/YA is a study of the children of all females who participated in the NLSY-79. The WLS is a study of graduates from high school in Wisconsin in 1957 (born 1937-1938) with a randomly selected sibling joining in 1977 or 1993. In each cohort, we selected sets of *full* siblings, restricting to +/- 5 year difference in birth years to improve the plausibility of the assumption of shared family background (in sensitivity analyses, we vary this restriction).

### Measures

#### Adult Body Mass Index

We calculated BMI by dividing weight by squared height (kg/m^2^). We focused on BMI measured at age 20-64 to reduce the risk of bias due to differential mortality rates. To remove the influence of outliers and biologically implausible values, we set height values less than 4.5 feet (1.37 meters) and more than 7 feet (2.13 metres) and BMI values outside the range of 13-70 to missing (approximately 0.1% of observations).

Adult height and weight were measured by self-report in each cohort. Adult height and weight were measured on multiple occasions in the NLSY-79, NLSY-97, and NLSY-79 C/YA and only once (1992-1994) in the WLS (see Supplementary Information for more detail). In cases where weight but not height was available at a given data collection, we used the last observed value for height, provided it was collected at age 20 or later, given the broad stability of height during adulthood [50].

#### Adolescent Cognitive ability

Cognitive ability was measured in each cohort using validated tests capturing multiple domains of cognition. In the NLSY-79 and NLSY-97 this was tested using the Armed Services Vocational Aptitude Battery (ASVAB), which participants sat in 1980 (age 15-23) and 1998-1999 (age 13-20), respectively. For both studies, and to ensure comparability with other cohorts, we used age-normed centile scores (range 0-100) provided with the data. These combine weighted scores for the mathematical knowledge, arithmetic reasoning, word knowledge, and paragraph comprehension subsections of the ASVAB (for the NLSY-79, this is the 2006 norming exercise). In sensitivity analyses, we converted cognitive ability centiles to z-scores instead.

In the NLSY-79 C/YA, cognitive ability was measured using three subscales from the Peabody Individual Achievement Test (PIAT), administered at ages 5–14 years (reading comprehension, reading recognition, and mathematical ability). Age-normed centile scores are available for each test, which we averaged. Given the PIAT was administered on multiple occasions, for age comparability with the NLSY-79 and NLSY-97, we used the last available measurement. Note, children were not eligible to complete the reading comprehension test if they obtained a low score on the reading recognition test, so we averaged the reading recognition and mathematical ability if only these were available [42]. Two- and three-test averaged PIAT scores were highly correlated (ρ = 0.97).

Cognitive ability was measured in the WLS using the Henmon–Nelson test, a group-administered, multiple-choice assessment containing verbal and quantitative items. The Henmon–Nelson test was sat by students in all Wisconsin high schools at varying school grades from the 1930s through the 1960s. We again used centile scores, with norming based on national test takers (range 0-100).

#### Socio-Economic Position

We included a measure of early socioeconomic position (SEP) to examine whether adjusting for this factor generates similar associations between cognitive ability and BMI *between families* as *within families*. For the NLSY-79, we used a composite measure of SEP [51] that has been used in multiple studies in the cognitive epidemiology literature [42,45], including a study examining the association between cognitive ability and BMI [26]. The measure averages z-scores for family income, parental occupational prestige, and mother’s and father’s years of education, each measured in 1978 or 1979. For the NLSY-79 CYA, we followed a similar approach and averaged z-scores for family income and mother’s education [42] (information on father’s education and on occupation is not available), using observations closest to a participant’s 18^th^ birthday. For the NLSY-97, we averaged z-scores for mother’s and father’s years of education and family income (occupational data not available), each of which were measured in 1997. For the WLS, we used a variable extracted from a factor analysis of family income (averaged between 1957-1960), parental occupational prestige, and mother’s and father’s years of education (recorded in 1957) that is supplied with the dataset. Note, in each cohort except the NLSY-79 CYA, the measure of SEP is fixed within a sibling set. More information on the SEP variables can be found in the Supplementary Information.

#### Covariates

Sibling designs do not control for confounding factors that vary among siblings. We included variables for cohort, sex, ethnic group (White, Black, Hispanic), age, birth order, and maternal age at birth. Other potential confounding variables were not measured consistently across cohorts, though data on childhood health were available in the NLSY-79 and NLSY-97, which we use in a robustness check (see below). Note, cohort and ethnic group are fixed within sibships and were included to reduce bias in between family associations. See Supplementary Information for more detail on the definitions of the covariates.

### Statistical Analysis

Our main analytical approach was to estimate linear random effects “within-between” (REWB) [52] models of the following form, pooling data from the individual cohorts:

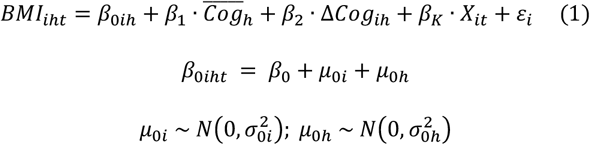

where *BMI*_*iht*_ is BMI for individual *i* from household *h* at time *t*; *β*_0*ih*_ is the intercept, comprising a fixed-effect (*β*_0_) and random intercepts at the individual (*μ*_0*i*_) and household (*μ*_0*h*_) level; 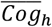 is the mean level of cognitive ability in a given household *h* (the between-family effect); and Δ*Cog*_*ih*_ is the sibling-specific deviation from the mean household cognitive ability (the within-family effect). *X*_*it*_ is a vector of control variables, specified above, with age, birth order and maternal age modelled with natural cubic splines (2 degrees of freedom each) [53] to account for potential non-linearities in their relationship with BMI [8]. To account for potential cross-cohort differences, we also included interaction terms between cohort and sex, ethnic group, age, and (depending on the model) socioeconomic position. We estimated models including and not including adjustment for SEP. Our interest was in the change in the coefficient *β*_1_ (between family association between cognitive ability and BMI) following adjustment for SEP and the relative size of the coefficient *β*_2_ (within family association).

The association between cognitive ability and BMI may differ markedly across the cohorts, so we repeated Model 1 for each cohort separately and excluding a single cohort, in turn (excluding individual random intercepts when analyzing the WLS on its own). We also repeated Model 1 including interactions between mean cognitive ability and cognitive ability deviations (Δ*Cog*_*ih*_) and (linear) age as the previous results suggest a strengthening association across the life course [26,31]. As a robustness check, we ran Model 1 including further control for childhood self-rated health (categories: excellent, very good, good, fair, poor) in the subset of cohorts with this data (NLSY-79 and NLSY-97).

Sibling designs can introduce bias through “carryover effects”, where one sibling influences another. Estimates may be attenuated if a sibling’s cognitive ability is important not just for their own BMI but their siblings’ BMI, too (for instance, through modelling a particular health behaviour such as smoking). To explore this possibility, we repeated Model 1 splitting the set of two sibling households according to whether the older or younger sibling had the higher cognitive ability score. On the assumption that older siblings will be more influential, within-family associations between cognitive ability and BMI should be smaller where the older sibling has the higher cognitive ability.

Next, we examined the association between cognitive ability and BMI across the distribution of BMI using residualized quantile regression (RQR) [47]. Unlike standard conditional quantile regression [54], RQR allows for the inclusion of (family) fixed effects in quantile regression models while retaining clear interpretability [55], a necessity here given we have more than one observation per family. RQR involves two steps: in the first step, the exposure variable (here, cognitive ability) is regressed upon control variables using linear regression (sex, age, ethnic group, birth order, family fixed effects) from which residuals are calculated. Where control variables have been chosen satisfactorily, these residuals represent exogenous variation in the exposure which can be used to estimate causal effects. In the second step, the outcome variable (here, BMI) is then regressed upon the residuals using quantile regression to obtain estimates of the effect of the exposure upon the unconditional outcome *distribution*. Bootstrapping is used to calculate standard errors and confidence intervals. Here, we used 500 (cluster-robust) bootstraps, and produced quantile regression estimates for each decile (10^th^, 20^th^, …, 90^th^ centiles) of the BMI distribution. As our measure of cognitive ability was time-invariant, we only used one observation per individual in the RQR procedure. In the main analysis, we used the first observation per individual, but also repeated the analysis using participants’ last observations. To obtain a comparable RQR estimate of the between-family association, we estimated RQR models using data from a single (randomly selected) individual in each household, dropping the household fixed effects from the first stage regression.

All analyses were performed in R version 4.12 [56]. Complete-case data were used in each analysis, given the difficulty accounting for the complex, multilevel structure of the data in multiple imputation models. Robust regression was used to absorb the household fixed effects in the RQR first stage regression.

## Results

### Descriptive Statistics

We identified 20,889 individuals (9,726 households) who were part of sibling sets. Of these, 14,560 individuals (6,665 households) had valid data for adult BMI for two or more siblings. Approximately 16% of individuals were excluded due to missing cognitive ability or covariate data, or lack of discordance in cognitive ability by siblings (N = 28). The analytical sample size was therefore 12,250 siblings from 5,602 households (118,355 observations), 59% of the adult sibling sample. Tables S1 and S2 detail sample sizes and attrition rates for each cohort.

Descriptive statistics for time-invariant characteristics are displayed in Table 1. Figure S2 shows the distribution of cognitive ability in each cohort. Cognitive ability levels were close to the population mean in the NLSY-79 and NLSY-97 Main samples and the NLSY-79 YCA. The NLSY-79 and NLSY-97 Oversamples were below population means, while cognitive ability levels in the WLS were above population means. While there was less variation in cognitive ability scores within than between families, variability was still substantial; the within family SD for cognitive ability was between 50-70% of the between family SD in each cohort. Overall, values for sex, age, ethnic group, and birth order were similar among those who had above (family-specific) average cognitive ability and below (family-specific) average cognitive ability (see Table 1, Table S3, and Figure S3). However, there was some evidence that siblings with above (family-specific) average cognitive ability had better childhood health (Figure S4).

**Table 1:** Descriptive statistics for time-invariant variables by cognitive ability (relative to family average) in four cohort studies. Mean (SD) and N (%).

| Variable | Group, cognitive ability | NLSY-79 Main | NLSY-79 Oversample | NLSY-79 CYA | NLSY-97 Main | NLSY-97 Oversample | WLS |
| --- | --- | --- | --- | --- | --- | --- | --- |
| N | Total Sample | 2,556 | 1,755 | 2,809 | 1,873 | 628 | 2,629 |
|  | Below Average | 1,276 (49.9%) | 897 (51.1%) | 1,396 (49.7%) | 936 (50%) | 319 (50.8%) | 1,314 (50%) |
|  | Above Average | 1,280 (50.1%) | 858 (48.9%) | 1,413 (50.3%) | 937 (50%) | 309 (49.2%) | 1,315 (50%) |
| Observations | Total Sample | 45,842 | 27,332 | 13,954 | 21,332 | 7,266 | 2,629 |
|  | Below Average | 22,886 (49.9%) | 14,041 (51.4%) | 6,894 (49.4%) | 10,507 (49.3%) | 3,668 (50.5%) | 1,314 (50%) |
|  | Above Average | 22,956 (50.1%) | 13,291 (48.6%) | 7,060 (50.6%) | 10,825 (50.7%) | 3,598 (49.5%) | 1,315 (50%) |
| Households | Total Sample | 1,113 | 750 | 1,246 | 889 | 291 | 1,313 |
|  | Below Average | 1,113 (100%) | 750 (100%) | 1,246 (100%) | 889 (100%) | 291 (100%) | 1,313 (100%) |
|  | Above Average | 1,113 (100%) | 750 (100%) | 1,246 (100%) | 889 (100%) | 291 (100%) | 1,313 (100%) |
| Cognitive Ability | Total Sample | 50.03 (29.69) | 27.93 (22.85) | 51.27 (24.84) | 50.6 (28.97) | 26.56 (22.23) | 62.67 (25.15) |
|  | Below Average | 38.91 (27.48) | 18.59 (17.68) | 41.68 (23.63) | 39.67 (26.45) | 17.4 (17.1) | 51.39 (24.42) |
|  | Above Average | 61.11 (27.6) | 37.69 (23.56) | 60.74 (22.24) | 61.52 (27.22) | 36.01 (22.96) | 73.95 (20.35) |
| Female | Total Sample | 1,255 (49.1%) | 833 (47.46%) | 1,410 (50.2%) | 916 (48.91%) | 302 (48.09%) | 1,378 (52.42%) |
|  | Below Average | 620 (48.59%) | 426 (47.49%) | 725 (51.93%) | 435 (46.47%) | 146 (45.77%) | 678 (51.6%) |
|  | Above Average | 635 (49.61%) | 407 (47.44%) | 685 (48.48%) | 481 (51.33%) | 156 (50.49%) | 700 (53.23%) |
| Male | Total Sample | 1,301 (50.9%) | 922 (52.54%) | 1,399 (49.8%) | 957 (51.09%) | 326 (51.91%) | 1,251 (47.58%) |
|  | Below Average | 656 (51.41%) | 471 (52.51%) | 671 (48.07%) | 501 (53.53%) | 173 (54.23%) | 636 (48.4%) |
|  | Above Average | 645 (50.39%) | 451 (52.56%) | 728 (51.52%) | 456 (48.67%) | 153 (49.51%) | 615 (46.77%) |
| Birth Order | Total Sample | 2.98 (1.8) | 3.74 (2.33) | 1.94 (0.96) | 1.97 (1) | 2.26 (1.12) | 2.19 (1.31) |
|  | Below Average | 3 (1.83) | 3.69 (2.28) | 1.97 (0.94) | 2.07 (0.98) | 2.27 (1.09) | 2.21 (1.29) |
|  | Above Average | 2.95 (1.77) | 3.78 (2.37) | 1.9 (0.97) | 1.88 (1) | 2.25 (1.15) | 2.18 (1.32) |
| Maternal Age | Total Sample | 25.82 (5.71) | 25.88 (6.34) | 24.58 (4.9) | 25.45 (4.71) | 24.41 (5.12) | 27.32 (5.16) |
|  | Below Average | 25.81 (5.79) | 25.69 (6.34) | 24.68 (4.92) | 25.67 (4.75) | 24.42 (5.07) | 27.28 (5.17) |
|  | Above Average | 25.83 (5.63) | 26.08 (6.33) | 24.49 (4.88) | 25.23 (4.67) | 24.39 (5.17) | 27.37 (5.15) |

### Associations Between Adolescent Cognitive Ability and Adult BMI

In between-family analysis, higher adolescent cognition was associated with lower adult BMI; the difference in BMI moving from the lowest to highest percentile of cognition was −1.89 kg/m^2^ (95% CI = −2.41, −1.37; Figure 1). Adjusting for SEP attenuated the association to −1.23 kg/m^2^ (−1.79, −0.66). However, in *within-family* analysis the association was substantially weaker and confidence intervals crossed the null: a −0.13 kg/m^2^ (−0.70, 0.45) difference, approximately 90% smaller than the adjusted between-family association. A weak within family association between cognitive ability and BMI was found when conducted in each cohort separately, with confidence intervals overlapping the null in all cohorts (Figure 1). Expressed alternatively, for a person of average adult height (1.68m), a −0.13 kg/m^2^ difference in BMI is equivalent to a 0.36 kg lower weight.

**Figure 1:**
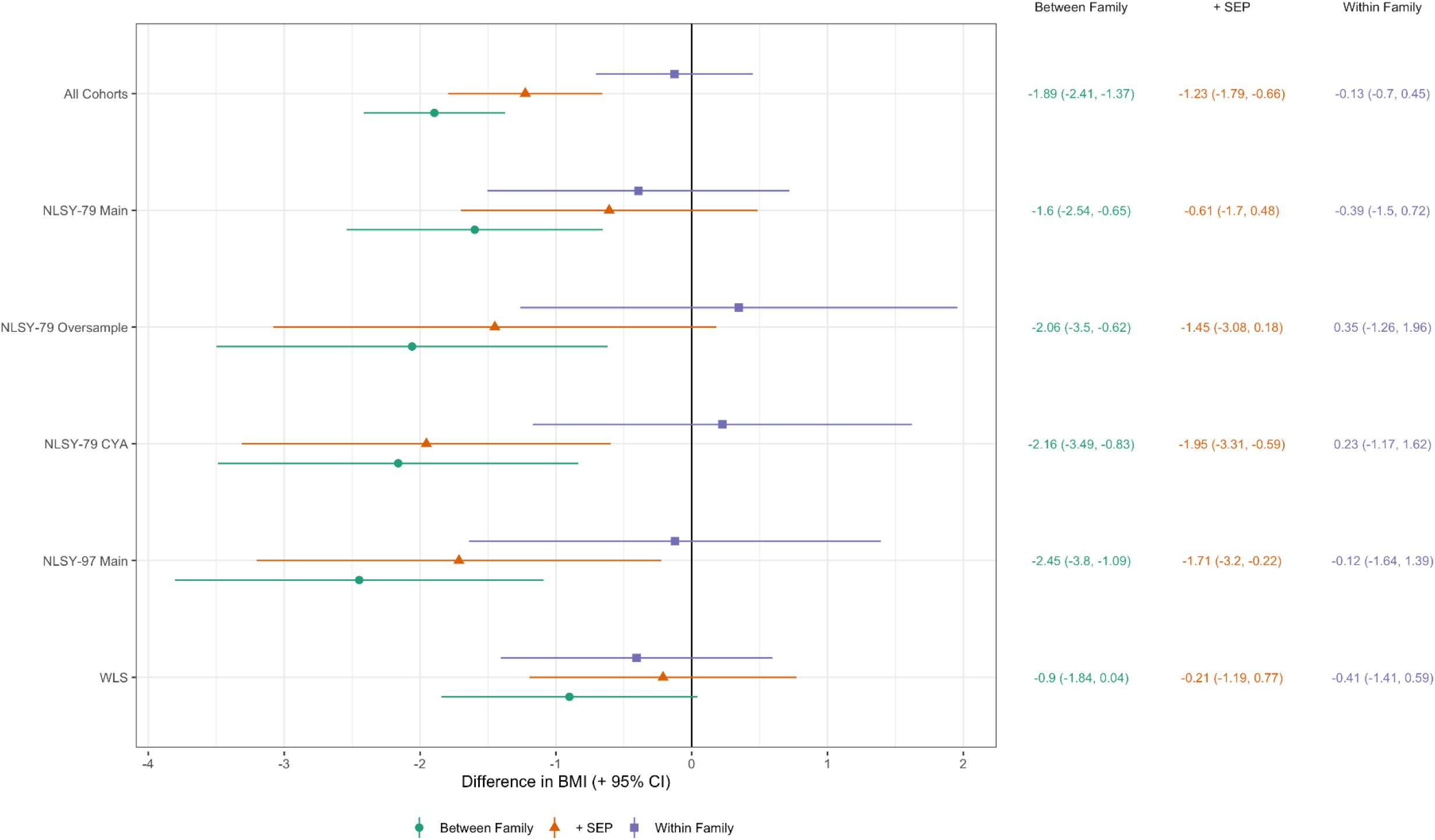
Between and within family associations between cognitive ability (centile rank) and BMI (+ 95% CIs). Estimates show predicted difference in BMI (kg/m^2^) comparing individual at the lowest to highest cognition (0^th^ to 100^th^ percentile). Derived from linear mixed effects models with random intercepts at household and individual level and age (two natural cubic splines), sex, cohort, birth order, and maternal age included as control variables.

Results examining age differences in the association between cognitive ability and BMI also showed a similar difference in between and within-family results (Figure 2). In *between family* analysis, the association was substantially stronger at older ages, rising from −0.81 kg/m^2^ (−1.38, −0.23) at age 20 to −1.91 kg/m^2^ (−2.50, −1.32) at age 60. The within family association, however, showed little change with age and was again small with confidence intervals crossing the null (Figure 2).

**Figure 2:**
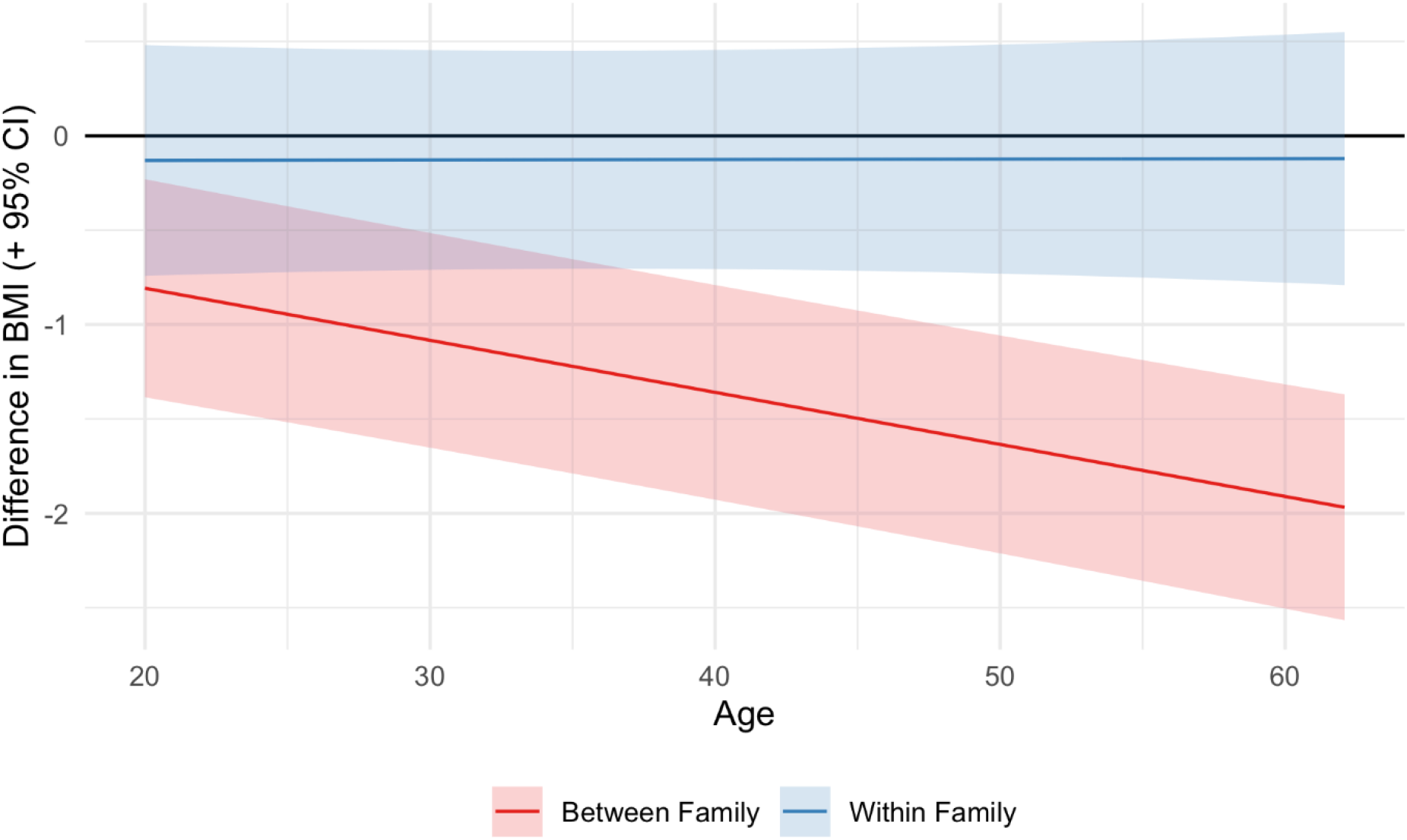
Between and within family associations between cognitive ability (percentile) and BMI by age. Derived from linear mixed effects models with random intercepts at household and individual level and age (two natural cubic splines), sex, cohort, birth order, maternal age, and SEP included as control variables. The lines show, at a given age, the predicted difference in BMI moving from bottom to highest cognitive ability centiles.

Finally, analysis of the association between cognitive ability and BMI, across the distribution of BMI, showed a similar pattern of results. In between-family analysis, cognitive ability was associated with lower BMI across the BMI distribution, with effect sizes largest at higher quantiles: the effect size at the was −0.15 kg/m^2^ (−0.49, 0.21) at the 10^th^ percentile −2.24 kg/m^2^ (−3.66, −0.81) at the 90^th^ percentile; Figure 3. In within-family analysis, effect sizes were smaller and confidence intervals crossed the null (90^th^ centile = −0.29 kg/m^2^; 95% CI = −1.88, 1.05).

**Figure 3:**
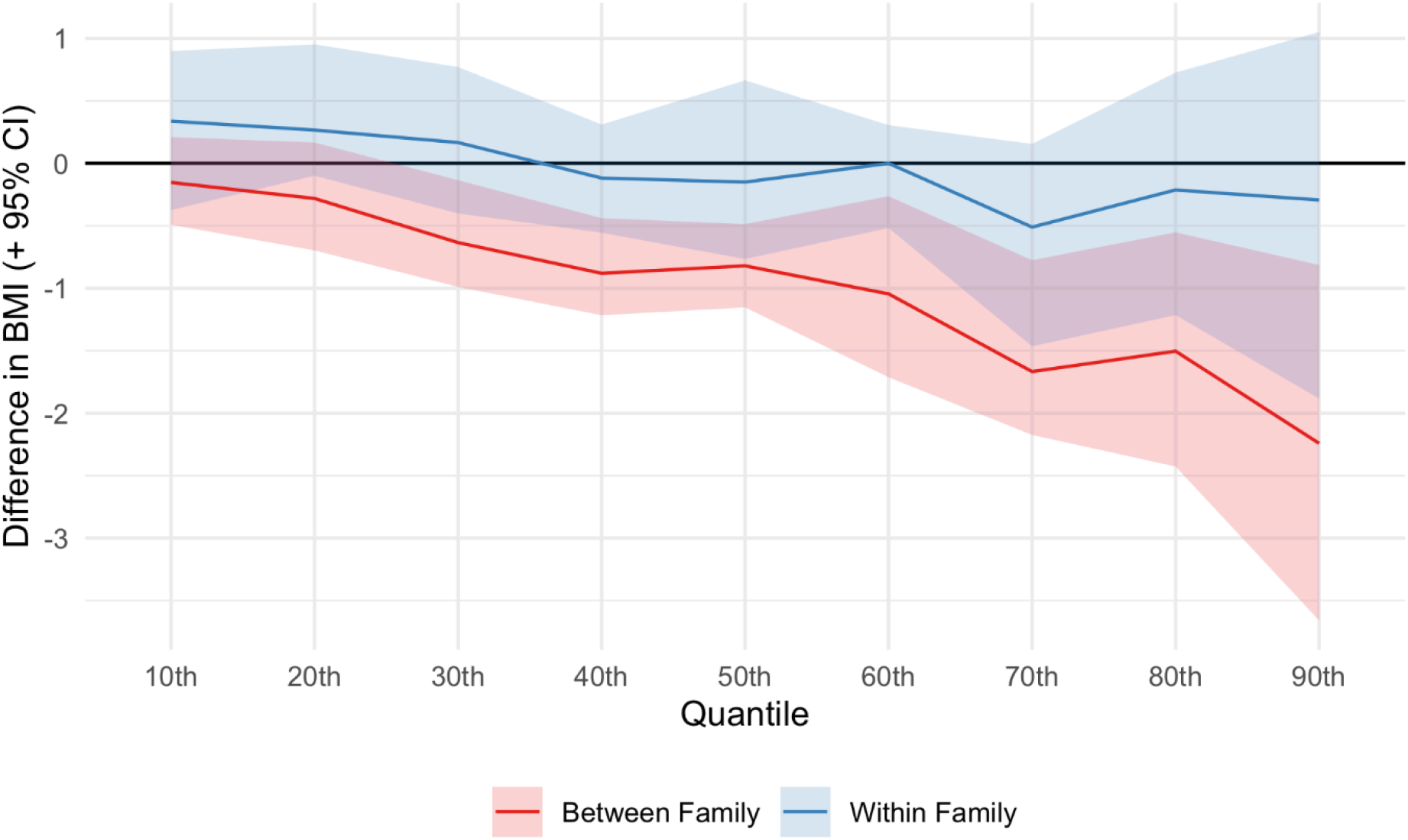
Associations between cognitive ability (percentile) and BMI by quantile of BMI. Derived from residualized quantile regressions using the first observation per individual (within family effect) and one (first) observation per household (between family effect). Age, maternal age, birth order,, sex, SEP, and cohort included as control variables in the first stage regressions with household fixed effects also included for within family models. Confidence intervals calculated using cluster-robust bootstrapping (percentile method, 500 replications). The estimates show, the predicted difference in BMI moving from bottom to highest cognitive ability centiles at a given centile of BMI – i.e., the results for the 50^th^ centile show the predicted difference in the median BMI among persons with highest cognitive ability compared with the median centile among individuals with lowest cognitive ability).

### Sensitivity Analyses

Qualitatively similar results were observed when varying the maximum age range to select sibling sets between 1-10 years (Figure S5), when excluding a single cohort from REWB models (Figure S6), and when using cognitive ability z-scores rather than ranks (Figure S7). Within-family estimates in the NLSY-79 and NLSY-97 combined were attenuated still further when including adolescent self-rated as a control variable. Consistent with a carryover effect, the within family association was weaker among two sibling families where the older sibling had the higher cognitive ability score, though confidence intervals had considerable overlap (Figure S8). Within-family associations between cognitive ability and the distribution of BMI generally remained weak when the last observation was used in RQR models (Figure S9).

## Discussion

### Main Findings

Using sibling data from four cohort studies, we found a sizeable difference in the between- and within-family estimates between adolescent cognitive ability and adult BMI. While higher cognitive ability was associated with lower BMI in between-family analysis, effect sizes in within-family analysis were small and confidence intervals overlapped the null. This difference was found across multiple specifications, including analyses conducted in separate cohorts, examining age-differences in association, and examining differences in association across the distribution of BMI.

### Explanation of Findings

Our results are consistent with the hypothesis that the association between adolescent cognitive ability and adult BMI observed in this and other studies [26–39] is substantially biased by factors which are shared by siblings. This finding is also consistent with recent work showing a marked attenuation in the genetic correlation between cognitive ability and BMI when using sibling data [57]. One plausible confounder that is shared by siblings is childhood socioeconomic position, which is in turn is associated with both adolescent cognition [58] and adult BMI [59,60]. Consistent with this, adjustment for childhood SEP attenuated associations in between-family analysis. However, SEP is a multidimensional construct typically captured with measurement error, leading to possible residual confounding. For instance, years of education does not capture quality of schooling and household income does not capture wealth. Besides SEP, other possible confounders shared between siblings include parental cognitive ability [42] and early neighbourhood food environments.

While our within-family estimates were still consistent with cognitive ability having a causal effect on BMI, the small effect sizes we identified are surprising given theory and previous results that health literacy and SEP (which cognitive ability should operate through) are important predictors of obesity, as well as other results in the wider cognitive epidemiology literature showing little attenuation when using sibling designs for other outcomes [61–63]. We note, however, that at a macro level, policies focusing on teaching the public about food choices appear to have little evidence of impact [5]. Further, quasi-experimental studies of compulsory schooling reforms – which are associated with small increases in cognitive ability [64] – and natural experiment studies of income windfalls find inconclusive (and sometimes conflicting) effects on obesity overall [65,66]. One reason for the small effect sizes found here may be that non-volitional factors, such as appetite and dietary norms, are of greater importance than conscious, reflective decision making for eating and exercise behaviour.

Another possibility is that our within-family estimates are biased and overly conservative. Sibling designs can introduce bias through three channels: measurement error of the exposure, confounding via non-shared factors, and carryover effects [46,67,68]. Measurement error is an unlikely explanation here as the cognitive tests used have high reliability [69,70]. Regarding non-shared factors, it is unclear which would bias the association downwards to such an extent – many candidates would be anticipated to upwardly bias the association or are otherwise relatively rare at a population level. In our data, adjusting for childhood self-reported health attenuated estimates still further. Childhood illnesses causing lifelong wasting are unlikely to be sufficiently common. We did, however, find suggestive and indirect evidence of carryover effects – associations were weaker when the older sibling had higher cognition. However, the analysis was low powered and point estimates continued to show relatively small effect sizes. One potentially source of carryover effects that was not explored in our analyses was differences in parental investments: there is evidence that parents compensate behaviourally for differences in siblings’ polygenic predisposition to high cognitive ability, at least in some families [71].

### Strengths and Limitations

Strengths of this study included the use of multiple samples and the longitudinal design: measures of cognitive ability were measured years before BMI, and we were able to increase statistical precision by including repeat measurement of BMI for each individual. Limitations of this study included a reliance on self-reported height and weight measurements, which likely contain measurement error [72]. Further, the degree of this error could be feasibly related to cognitive ability level. As noted, while sibling designs account for *shared* characteristics, non-shared factors may still bias associations (e.g. childhood infections or disease). There was also substantial attrition in the cohorts and, by using complete case data, there is thus a potential for selection bias. However, the number of successful follow-ups were similar among above and below (family) average cognitive ability groups.

## Conclusions

Associations between high adolescent cognitive ability and low adult BMI were partly attenuated by family SEP in between-family analysis, and substantially attenuated toward the null in within-family analysis. Our results are consistent with the hypothesis that the well-replicated association between high adolescent cognitive ability and low adult BMI is biased by factors such as SEP which are shared by siblings. Since such factors may confound associations between cognitive ability and other health outcomes, further research is required to test whether other results in the cognitive epidemiology literature are biased by family level SEP.

## Data Availability

Data are publicly available via the NLS Investigator (https://www.nlsinfo.org/investigator) and Wisconsin Longitudinal Study (https://www.ssc.wisc.edu/wlsresearch/) websites. Code to replicate the analysis (including NLSY variable tagsets) is available at https://osf.io/b4anm/.

https://www.nlsinfo.org/investigator

## Statements

### Author contributions

All authors conceived of and designed the study. LW conducted the analysis and wrote the first draft of the manuscript. DB and NMD provided comments and revisions. All authors read and approved the final manuscript.

## References

1 Ng M, Fleming T, Robinson M, et al. Global, regional, and national prevalence of overweight and obesity in children and adults during 1980–2013: a systematic analysis for the Global Burden of Disease Study 2013. The Lancet 2014;384:766–81. doi:10.1016/S0140-6736(14)60460-8

2 Tremmel M, Gerdtham U-G, Nilsson PM, et al. Economic Burden of Obesity: A Systematic Literature Review. International Journal of Environmental Research and Public Health 2017;14:435. doi:10.3390/ijerph14040435

3 McAllister EJ, Dhurandhar NV, Keith SW, et al. Ten Putative Contributors to the Obesity Epidemic. Critical Reviews in Food Science and Nutrition 2009;49:868–913. doi:10.1080/10408390903372599

4 Keith SW, Redden DT, Katzmarzyk PT, et al. Putative contributors to the secular increase in obesity: exploring the roads less traveled. Int J Obes 2006;30:1585–94. doi:10.1038/sj.ijo.0803326

5 Theis DR z., White M. Is Obesity Policy in England Fit for Purpose? Analysis of Government Strategies and Policies, 1992–2020. The Milbank Quarterly 2021;99:126–70. doi:10.1111/1468-0009.12498

6 WHO Regional Office for Europe. WHO European Regional Obesity Report 2022. 2022. https://apps.who.int/iris/handle/10665/353747 (accessed 13 May 2022).

7 Flegal KM, Carroll MD, Kit BK, et al. Prevalence of Obesity and Trends in the Distribution of Body Mass Index Among US Adults, 1999-2010. JAMA 2012;307:491–7. doi:10.1001/jama.2012.39

8 Johnson W, Li L, Kuh D, et al. How Has the Age-Related Process of Overweight or Obesity Development Changed over Time? Co-ordinated Analyses of Individual Participant Data from Five United Kingdom Birth Cohorts. PLoS Med 2015;12:e1001828.

9 NCD Risk Factor Collaboration. Trends in adult body-mass index in 200 countries from 1975 to 2014: a pooled analysis of 1698 population-based measurement studies with 19.2 million participants. The Lancet 2016;387:1377–96.

10 NCD Risk Factor Collaboration (NCD-RisC). Heterogeneous contributions of change in population distribution of body mass index to change in obesity and underweight. eLife 2021;10:e60060. doi:10.7554/eLife.60060

11 Majer IM, Mackenbach JP, van Baal PHM. Time trends and forecasts of body mass index from repeated cross-sectional data: a different approach. Stat Med 2013;32:1561–71. doi:10.1002/sim.5558

12 Jackson SE, Llewellyn CH, Smith L. The obesity epidemic – Nature via nurture: A narrative review of high-income countries. SAGE Open Medicine 2020;8:2050312120918265. doi:10.1177/2050312120918265

13 Elks CE, den Hoed M, Zhao JH, et al. Variability in the Heritability of Body Mass Index: A Systematic Review and Meta-Regression. Front Endocrin 2012;3. doi:10.3389/fendo.2012.00029

14 Deary IJ, Batty GD. Cognitive epidemiology. Journal of Epidemiology & Community Health 2007;61:378–84. doi:10.1136/jech.2005.039206

15 Deary IJ, Hill WD, Gale CR. Intelligence, health and death. Nat Hum Behav 2021;5:416–30. doi:10.1038/s41562-021-01078-9

16 Gottfredson LS. Intelligence: Is It the Epidemiologists’ Elusive ‘Fundamental Cause’ of Social Class Inequalities in Health? Journal of Personality and Social Psychology 2004;86:174–99. doi:10.1037/0022-3514.86.1.174

17 Calvin CM, Batty GD, Deary IJ. Cognitive Epidemiology: Concepts, Evidence, and Future Directions. In: Chamorro-Premuzic T, von Stumm S, Furnham A, eds. The Wiley-Blackwell Handbook of Individual Differences. Oxford, UK: : Wiley-Blackwell 2013. 427–60. doi:10.1002/9781444343120.ch16

18 Strenze T. Intelligence and socioeconomic success: A meta-analytic review of longitudinal research. Intelligence 2007;35:401–26. doi:10.1016/j.intell.2006.09.004

19 Grunert KG, Wills JM, Fernández-Celemín L. Nutrition knowledge, and use and understanding of nutrition information on food labels among consumers in the UK. Appetite 2010;55:177–89. doi:10.1016/j.appet.2010.05.045

20 Paasche-Orlow MK, Parker RM, Gazmararian JA, et al. The Prevalence of Limited Health Literacy. Journal of General Internal Medicine 2005;20:175–84. doi:10.1111/j.1525-1497.2005.40245.x

21 Cowburn G, Stockley L. Consumer understanding and use of nutrition labelling: a systematic review. Public Health Nutrition 2005;8:21–8. doi:10.1079/PHN2004666

22 Boylan S, Louie JCY, Gill TP. Consumer response to healthy eating, physical activity and weight-related recommendations: a systematic review. Obes Rev 2012;13:606–17. doi:10.1111/j.1467-789X.2012.00989.x

23 Reeve CL, Basalik D. Is health literacy an example of construct proliferation? A conceptual and empirical evaluation of its redundancy with general cognitive ability. Intelligence 2014;44:93–102. doi:10.1016/j.intell.2014.03.004

24 Yu ZB, Han SP, Cao XG, et al. Intelligence in relation to obesity: a systematic review and meta-analysis. Obes Rev 2010;11:656–70. doi:10.1111/j.1467-789X.2009.00656.x

25 Kanazawa S. Intelligence and obesity: which way does the causal direction go? Current Opinion in Endocrinology, Diabetes and Obesity 2014;21:339–44. doi:10.1097/MED.0000000000000091

26 Altschul DM, Wraw C, Gale CR, et al. How youth cognitive and sociodemographic factors relate to the development of overweight and obesity in the UK and the USA: a prospective cross-cohort study of the National Child Development Study and National Longitudinal Study of Youth 1979. BMJ Open 2019;9:e033011. doi:10.1136/bmjopen-2019-033011

27 Batty GD, Deary IJ, Macintyre S. Childhood IQ in relation to risk factors for premature mortality in middle-aged persons: the Aberdeen Children of the 1950s study. J Epidemiol Community Health 2007;61:241–7. doi:10.1136/jech.2006.048215

28 Batty GD, Deary IJ, Schoon I, et al. Mental ability across childhood in relation to risk factors for premature mortality in adult life: the 1970 British Cohort Study. Journal of Epidemiology & Community Health 2007;61:997–1003. doi:10.1136/jech.2006.054494

29 Batty GD, Gale CR, Mortensen LH, et al. Pre-morbid intelligence, the metabolic syndrome and mortality: the Vietnam Experience Study. Diabetologia 2008;51:436–43. doi:10.1007/s00125-007-0908-5

30 Belsky DW, Caspi A, Goldman-Mellor S, et al. Is Obesity Associated With a Decline in Intelligence Quotient During the First Half of the Life Course? Am J Epidemiol 2013;178:1461–8. doi:10.1093/aje/kwt135

31 Chandola T, Deary IJ, Blane D, et al. Childhood IQ in relation to obesity and weight gain in adult life: the National Child Development (1958) Study. Int J Obes 2006;30:1422–32. doi:10.1038/sj.ijo.0803279

32 Dahl Aslan AK, Starr JM, Pattie A, et al. Cognitive consequences of overweight and obesity in the ninth decade of life? Age and Ageing 2015;44:59–65. doi:10.1093/ageing/afu108

33 Hagger-Johnson G, Mõttus R, Craig LCA, et al. Pathways from childhood intelligence and socioeconomic status to late-life cardiovascular disease risk. Health Psychol 2012;31:403–12. doi:10.1037/a0026775

34 Halkjær J, Holst C, Sørensen TIA. Intelligence Test Score and Educational Level in Relation to BMI Changes and Obesity. Obesity Research 2003;11:1238–45. doi:10.1038/oby.2003.170

35 Kanazawa S. Childhood intelligence and adult obesity. Obesity 2013;21:434–40. doi:10.1002/oby.20018

36 Kumpulainen SM, Heinonen K, Salonen MK, et al. Childhood cognitive ability and body composition in adulthood. Nutr & Diabetes 2016;6:e223–e223. doi:10.1038/nutd.2016.30

37 Modig K, Bergman LR. Associations between intelligence in adolescence and indicators of health and health behaviors in midlife in a cohort of Swedish women. Intelligence 2012;40:82–90. doi:10.1016/j.intell.2012.02.002

38 Rosenblad A, Nilsson G, Leppert J. Intelligence level in late adolescence is inversely associated with BMI change during 22 years of follow-up: results from the WICTORY study. Eur J Epidemiol 2012;27:647–55. doi:10.1007/s10654-012-9713-7

39 Wimmelmann CL, Grønkjær M, Mortensen EL. Changes in BMI from young adulthood to late midlife in 1536 Danish men: The influence of intelligence and education. Obesity Medicine 2021;23:100334. doi:10.1016/j.obmed.2021.100334

40 Davies NM, Hill WD, Anderson EL, et al. Multivariable two-sample Mendelian randomization estimates of the effects of intelligence and education on health. eLife 2019;8:e43990. doi:10.7554/eLife.43990

41 Norris T, Bann D, Hardy R, et al. Socioeconomic inequalities in childhood-to-adulthood BMI tracking in three British birth cohorts. Int J Obes 2020;44:388–98. doi:10.1038/s41366-019-0387-z

42 Wraw C, Deary IJ, Der G, et al. Maternal and offspring intelligence in relation to BMI across childhood and adolescence. Int J Obes 2018;42:1610–20. doi:10.1038/s41366-018-0009-1

43 El-Sayed AM, Scarborough P, Galea S. Unevenly distributed: a systematic review of the health literature about socioeconomic inequalities in adult obesity in the United Kingdom. BMC Public Health 2012;12:18. doi:10.1186/1471-2458-12-18

44 Bann D, Johnson W, Li L, et al. Socioeconomic Inequalities in Body Mass Index across Adulthood: Coordinated Analyses of Individual Participant Data from Three British Birth Cohort Studies Initiated in 1946, 1958 and 1970. PLoS Med 2017;14:e1002214. doi:10.1371/journal.pmed.1002214

45 Wraw C, Deary IJ, Gale CR, et al. Intelligence in youth and health at age 50. Intelligence 2015;53:23–32. doi:10.1016/j.intell.2015.08.001

46 Sjölander A, Frisell T, Öberg S. Sibling Comparison Studies. Annual Review of Statistics and Its Application 2022;9:ull. doi:10.1146/annurev-statistics-040120-024521

47 Borgen NT, Haupt A, Wiborg ØN. A New Framework for Estimation of Unconditional Quantile Treatment Effects: The Residualized Quantile Regression (RQR) Model. 2021. doi:10.31235/osf.io/42gcb

48 Rothstein DS, Carr D, Cooksey E. Cohort Profile: The National Longitudinal Survey of Youth 1979 (NLSY79). Int J Epidemiol 2019;48:22–22e. doi:10.1093/ije/dyy133

49 Michael RT, Pergamit MR. The National Longitudinal Survey of Youth, 1997 Cohort. The Journal of Human Resources 2001;36:628. doi:10.2307/3069636

50 Cline MG, Meredith KE, Boyer JT, et al. Decline of height with age in adults in a general population sample: estimating maximum height and distinguishing birth cohort effects from actual loss of stature with aging. Hum Biol 1989;61:415–25.

51 Herrnstein RJ, Murray CA. The bell curve: intelligence and class structure in American life. 1st Free Press pbk. ed. New York: : Simon & Schuster 1996.

52 Bell A, Fairbrother M, Jones K. Fixed and random effects models: making an informed choice. Qual Quant 2019;53:1051–74. doi:10.1007/s11135-018-0802-x

53 Perperoglou A, Sauerbrei W, Abrahamowicz M, et al. A review of spline function procedures in R. BMC Medical Research Methodology 2019;19:46. doi:10.1186/s12874-019-0666-3

54 Koenker R, Bassett G. Regression Quantiles. Econometrica 1978;46:33. doi:10.2307/1913643

55 Borgen NT, Haupt A, Wiborg ØN. Quantile Regression Estimands and Models: Revisiting the Motherhood Wage Penalty Debate. 2020. doi:10.31235/osf.io/9avrp

56 R Core Team. R: A Language and Environment for Statistical Computing. Vienna, Austria: : R Foundation for Statistical Computing 2020. https://www.R-project.org/

57 Howe LJ, Nivard MG, Morris TT, et al. Within-sibship genome-wide association analyses decrease bias in estimates of direct genetic effects. Nat Genet 2022;54:581–92. doi:10.1038/s41588-022-01062-7

58 Connelly R, Gayle V. An investigation of social class inequalities in general cognitive ability in two British birth cohorts. The British Journal of Sociology 2019;70:90–108. doi:10.1111/1468-4446.12343

59 Brisbois TD, Farmer AP, McCargar LJ. Early markers of adult obesity: a review. Obesity Reviews 2012;13:347–67. doi:10.1111/j.1467-789X.2011.00965.x

60 Senese LC, Almeida ND, Fath AK, et al. Associations Between Childhood Socioeconomic Position and Adulthood Obesity. Epidemiologic Reviews 2009;31:21–51. doi:10.1093/epirev/mxp006

61 Bratsberg B, Rogeberg O. Childhood socioeconomic status does not explain the IQ-mortality gradient. Intelligence 2017;62:148–54. doi:10.1016/j.intell.2017.04.002

62 Iveson MH, Čukić I, Der G, et al. Intelligence and all-cause mortality in the 6-Day Sample of the Scottish Mental Survey 1947 and their siblings: testing the contribution of family background. International Journal of Epidemiology 2018;47:89–96. doi:10.1093/ije/dyx168

63 Jokela M, Batty GD, Deary IJ, et al. Sibling Analysis of Adolescent Intelligence and Chronic Diseases in Older Adulthood. Annals of Epidemiology 2011;21:489–96. doi:10.1016/j.annepidem.2011.01.008

64 Ritchie SJ, Tucker-Drob EM. How Much Does Education Improve Intelligence? A Meta-Analysis. Psychol Sci 2018;29:1358–69. doi:10.1177/0956797618774253

65 Galama TJ, Lleras-Muney A, van Kippersluis H. The Effect of Education on Health and Mortality: A Review of Experimental and Quasi-Experimental Evidence. National Bureau of Economic Research 2018. doi:10.3386/w24225

66 Au N, Johnston DW. Too Much of a Good Thing? Exploring the Impact of Wealth on Weight. Health Econ 2015;24:1403–21. doi:10.1002/hec.3094

67 Sjölander A, Frisell T, Kuja-Halkola R, et al. Carryover Effects in Sibling Comparison Designs. Epidemiology 2016;27:852–8. doi:10.1097/EDE.0000000000000541

68 Frisell T, Öberg S, Kuja-Halkola R, et al. Sibling comparison designs: bias from non-shared confounders and measurement error. Epidemiology 2012;23:713–20. doi:10.1097/EDE.0b013e31825fa230

69 Herd P, Carr D, Roan C. Cohort Profile: Wisconsin longitudinal study (WLS). Int J Epidemiol 2014;43:34–41. doi:10.1093/ije/dys194

70 Garrison SM, Rodgers JL. Casting doubt on the causal link between intelligence and age at first intercourse: A cross-generational sibling comparison design using the NLSY. Intelligence 2016;59:139–56. doi:10.1016/j.intell.2016.08.008

71 Breinholt A, Conley D. Child-Driven Parenting: Differential Early Childhood Investment by Offspring Genotype. National Bureau of Economic Research 2020. doi:10.3386/w28217

72 Scholes S, Ng Fat L, Moody A, et al. Does the use of prediction equations to correct self-reported height and weight improve obesity prevalance estimates? A pooled cross-sectional analysis of Health Survey for England data. Epidemiology 2022. doi:10.1101/2022.01.28.22270014

