## Supplemental Information for "The Association Between Cognitive Ability and Body Mass Index: A Sibling-Comparison Analysis in Four Longitudinal Studies"

Supplementary Information

### Participants

The NLSY-79 (Rothstein et al., 2019) and NLSY-97 (Michael & Pergamit, 2001) are ongoing studies of young people that began in 1979 and 1997, respectively. 12,886 individuals aged 14-22 (born 1957-1964) were initially recruited to the NLSY-79, with participants followed annually until 1994 and biennially thereafter. 8,894 individuals aged 12-17 (born 1980-1984) were initially recruited to the NLSY-97. Participants were followed annually to 2011 and biennially thereafter. The latest (mainstage) survey data available for the studies are from 2018 and 2019, respectively (ages 53-62 and 34-40). Beside a nationally representative sample, the NLSY-79 and NLSY-97 also included an oversample of ethnic minority individuals, with the NLSY-79 further including oversamples of individuals from military or economically disadvantaged backgrounds. The military oversample was dropped in later years and is not included here.

Recruitment to the NLSY-79 and NLSY-97 took place at the household level (8,770 and 6,814 households, respectively), with all residents within the age brackets eligible for inclusion; both studies thus contain data from multiple sibling sets. Here, we use data from the NLSY Kinship Links (Rodgers et al., 2016) to determine natural siblings, restricting to +/- 5 year difference in birth years to improve the plausibility of the assumption of shared family background. (Results were qualitatively similar varying the age range between 1-10 years.) For each household, we apply this rule such that the maximum number of siblings from a household are retained. Where multiple solutions exist, we choose the set containing the oldest siblings. There are 4,934 and 3,563 such siblings in the NLSY-79 and NLSY-97, respectively (2,107 and 1,673 households). Given the differences in the main and oversample participants, in analyses we treat these as separate cohorts.

The NLSY-79 C/YA is an ongoing study following all natural children of mothers from the NLSY-79 sample (Rothstein et al., 2019). The study consists of biennial interviews with mothers and their children, with the latest data available from 2018. From 1994, children aged 15 or above were followed independently of their mothers and complete a “Young Adult” questionnaire that is similar in scope to the NLSY-79 questionnaire proper. 11,545 children have been included in at least one data collection. Given the recruitment procedure, the age range is wider than for the NLSY-79 or NLSY-97, including within families. We again use the NLSY Kinship Links (Rodgers et al., 2016) to identify natural siblings, restricting to +/- 5 year difference in birth years to construct sibling sets. The NLSY-79 C/YA contains data from 4,219 such siblings (1,859 households), though not all of these individuals had reached adulthood by the final follow-up.

The WLS (Herd et al., 2014) comprises 10,317 individuals who graduated from high school in Wisconsin in 1957 (born 1937-1938). Participants have been followed intermittently between 1957 and 2011 (age 74), with a selected sibling of the participant also followed from 1977 to 2011. Restricting to +/- 5 year difference in birth years, the WLS contains data from 8,295 siblings (4,142 households).

### Measures

#### Adult Body Mass Index

Adult height and weight were measured by self-report in each cohort. Adult height and weight were measured on multiple occasions in the NLSY-79, NLSY-97, and NLSY-79 C/YA and only once (1992-1994) in the WLS. In the NLSY-79, height was measured in 1981, 1982, 1985, and 2006-2018, while weight was measured in 1981, 1982, 1985, 1986, 1988-90, and 1992-2016. In the WLS, height and weight were measured in 1992 for original sample members and in 1994 for siblings.

#### Adolescent Socioeconomic Position

In line with multiple previous studies in the cognitive epidemiology literature (Altschul et al., 2019; Wraw et al., 2015, 2018), we measured adolescent socioeconomic position (SEP) using the composite variable defined in Herrnstein and Murray’s (1996, pp. 597–599) Bell Curve. This variable averages z-scores for family income (averaged across 1978 and 1979), parental occupational prestige, and mother’s and father’s years of education (measured in 1979). Where one or more of these component measures is missing (e.g. family income), the remaining measures are averaged instead (e.g. parental education and occupational prestige). To validate our variable, we compared it against the original variable which is downloadable from Eric Rasmusen’s (2007) *Bell Curve Page*. However, our variable was highly correlated with the original variable (ρ = 0.97) but had less missingness (n = 72 vs 810). This could be due to data cleaning in the NLSY-79 source files since the Bell Curve was released.

We constructed similar composite z-score measures of adolescent SEP for the NLSY-97 and NLSY-79 YCA, with the limitation that all measure available in the NLSY-79 were not available in these other studies. For the NLSY-79 CYA, we averaged z-scores for family income and mother’s education (information on father’s education and on occupation is not available) using observations closest to a participants 18^th^ birthday (and not after age 22, for consistency with the NLSY-79). This is similar to the measure of SEP used in another paper in the cognitive epidemiology literature (Wraw et al., 2018) though they used information from earlier in childhood. Note, the data on family income was obtained from the mothers as part of the NLSY-79 follow-ups.

For the NLSY-97, we averaged z-scores for mother’s and father’s years of education and family income (occupational data not available), each of which were measured in the first wave (1997). For the WLS, we use a variable extracted from a factor analysis of family income (averaged between 1957-1960), parental occupational prestige, and mother’s and father’s years of education (recorded in 1957) that is supplied with the dataset (ses57). Note, in each cohort except the NLSY-79 CYA, the measure of SEP is fixed within a sibling set.

#### Covariates

We included variables for cohort, sex, ethnic group (White, Black, Hispanic), age, birth order, and maternal age at birth. Due to low numbers of non-white participants, data on race is not available for the WLS, so we assumed all participants were white. Maternal age at birth was missing in a number of cases. In the NLSY-79, NLSY-97 and WLS, we overwrote using siblings’ values, provided these were not overly discordant (> 5 years [> 2 years for WLS] difference between implied maternal age from sibling responses). We set the maternal age variable to missing if it was outside the range 13-50.

Birth order was defined differently in each cohort. In the NLSY-79, we used an (undocumented) variable supplied with the data on number of older siblings. In the NLSY-79 CYA, birth order was defined as (natural mother) parity. In the NLSY-97, following Rohrer et al. (2015), birth order was defined by summing older resident siblings of any type (natural, half, step, foster siblings, etc.) and non-resident full siblings using household grid data from 1997. In the WLS, birth order was derived using data on siblings of any type, collected from graduates during the 1975 interview. We truncated the birth order variable at 10 to reduce the influence of outliers.

Childhood and adolescent self-rated health was captured in the NLSY-79 and the NLSY-97 only. In the NLSY-79, early self-rated health was measured with a retrospective question asked in 2012, 2014, or 2016: “Consider your health when you were growing up, from birth to age 17. Would you say your health during that time was excellent, very good, good, fair, or poor?” In the NLSY-97, adolescent self-rated health was measured prospectively in 1997 with the question “In general, how is your health? Excellent, very good, good, fair, poor”

### Figures

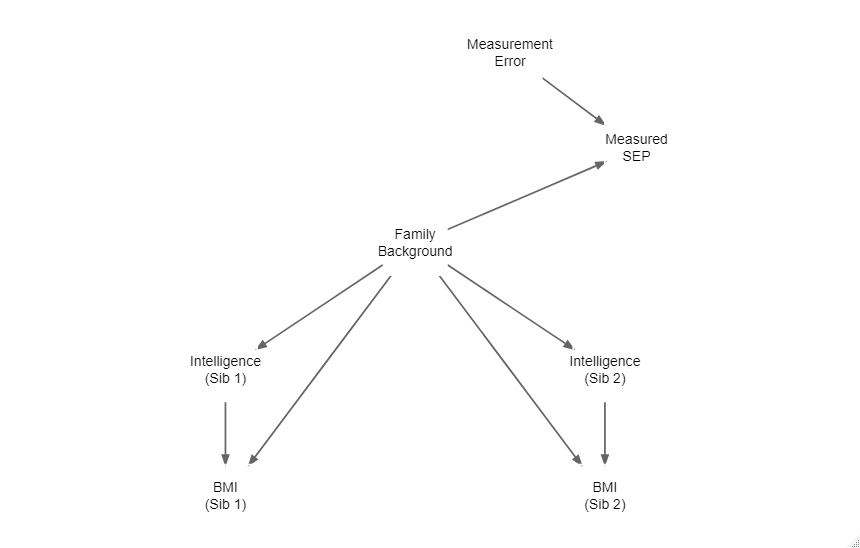

Figure S1: Directed Acyclic Graph. The association between (adolescent) cognitive ability and adult BMI may be confounded by family background (including dynastic genetic effects) and other factors shared between siblings. Existing studies typically attempt to control for family background with measured SEP, which because of the few high-level variables that are used, can be thought of measuring family background with some measurement error. Thus, controlling for measured SEP does not full block confounding through family background. The sibling design instead accounts for family background by design.

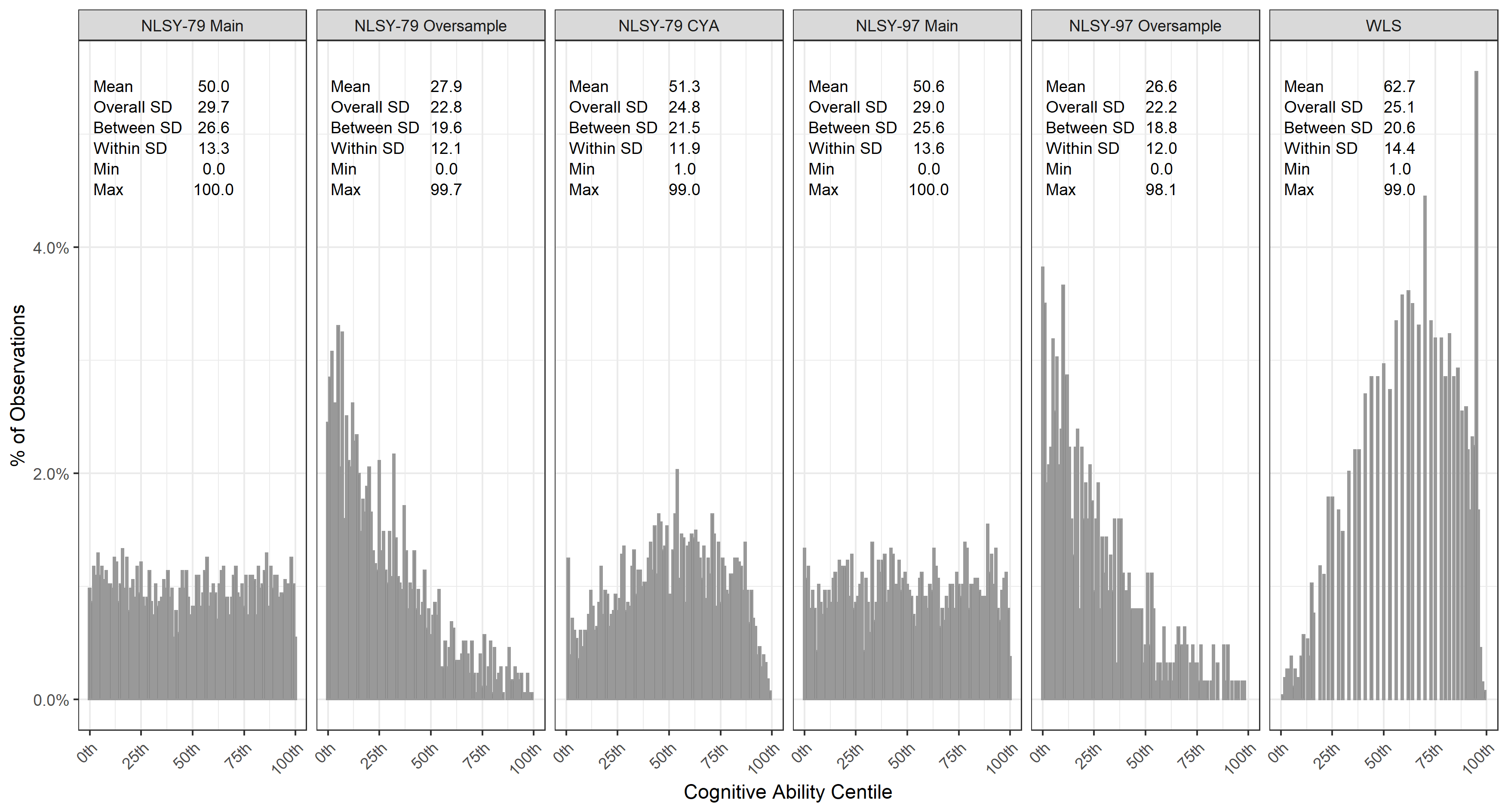

Figure S2: Descriptive statistics and distribution of cognitive ability (percentiles) by cohort.

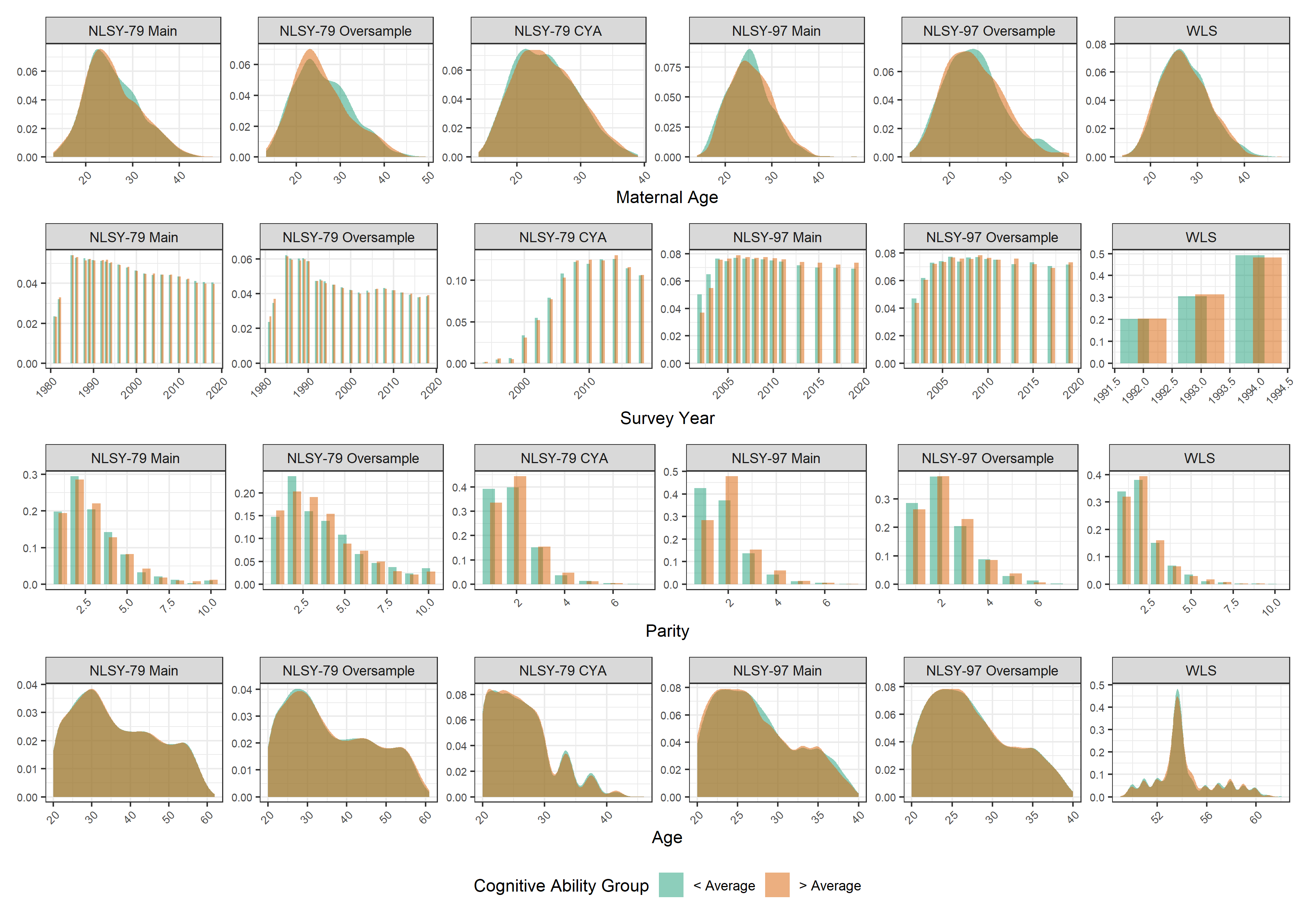

Figure S3: Distribution of control variables by (within-family) cognitive ability group.

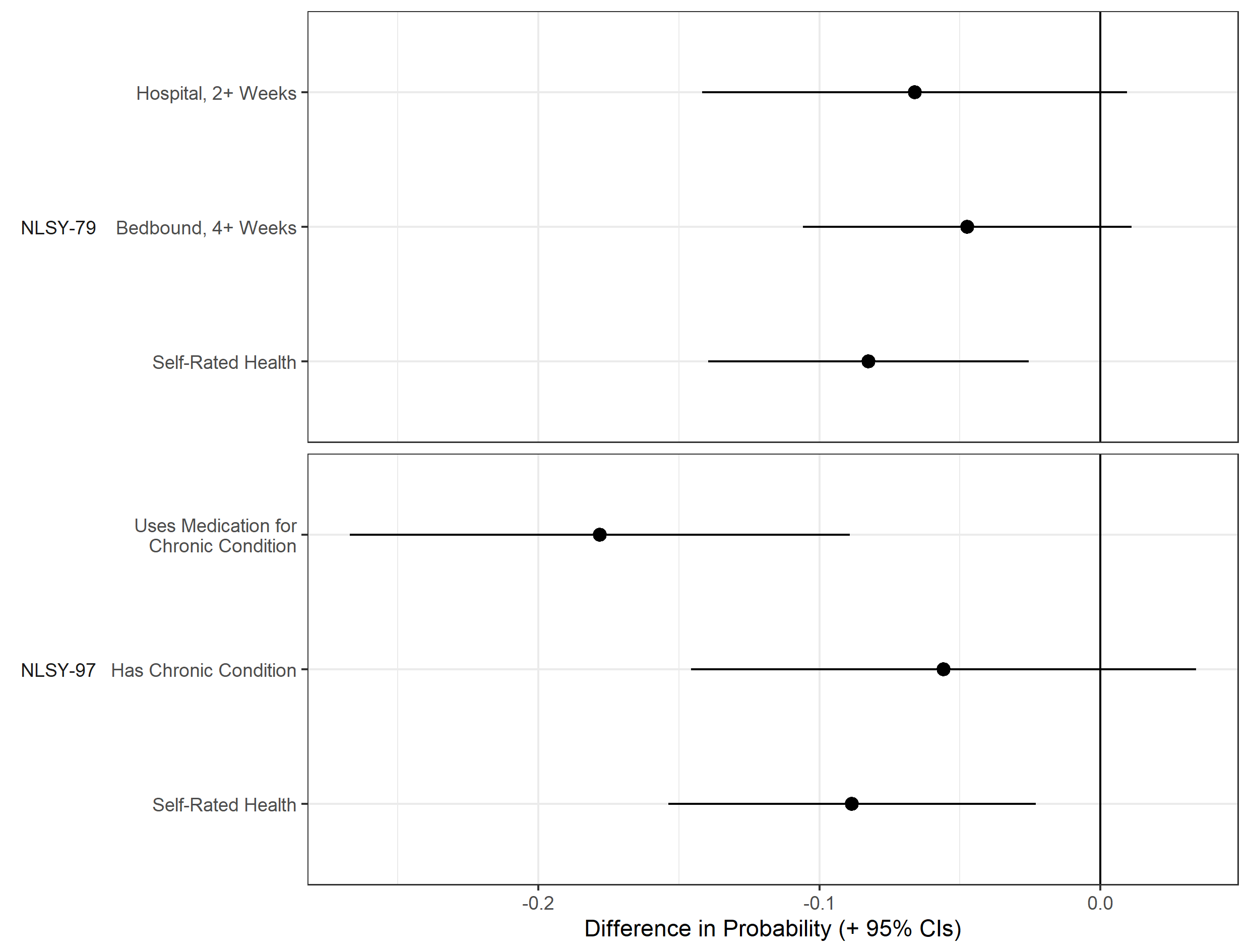

Figure S4: Association between (within-family) cognitive ability and probability of poor childhood health (+95% CI). Estimates drawn from linear probability fixed effects models. Childhood health variables from NLSY-79 and NLSY-97. Self-rated health converted to binary variable for this analysis (poor or fair vs good, very good or excellent)..Estimates show the difference in probability of each outcome as cognition centile increases (from 0 to 100).

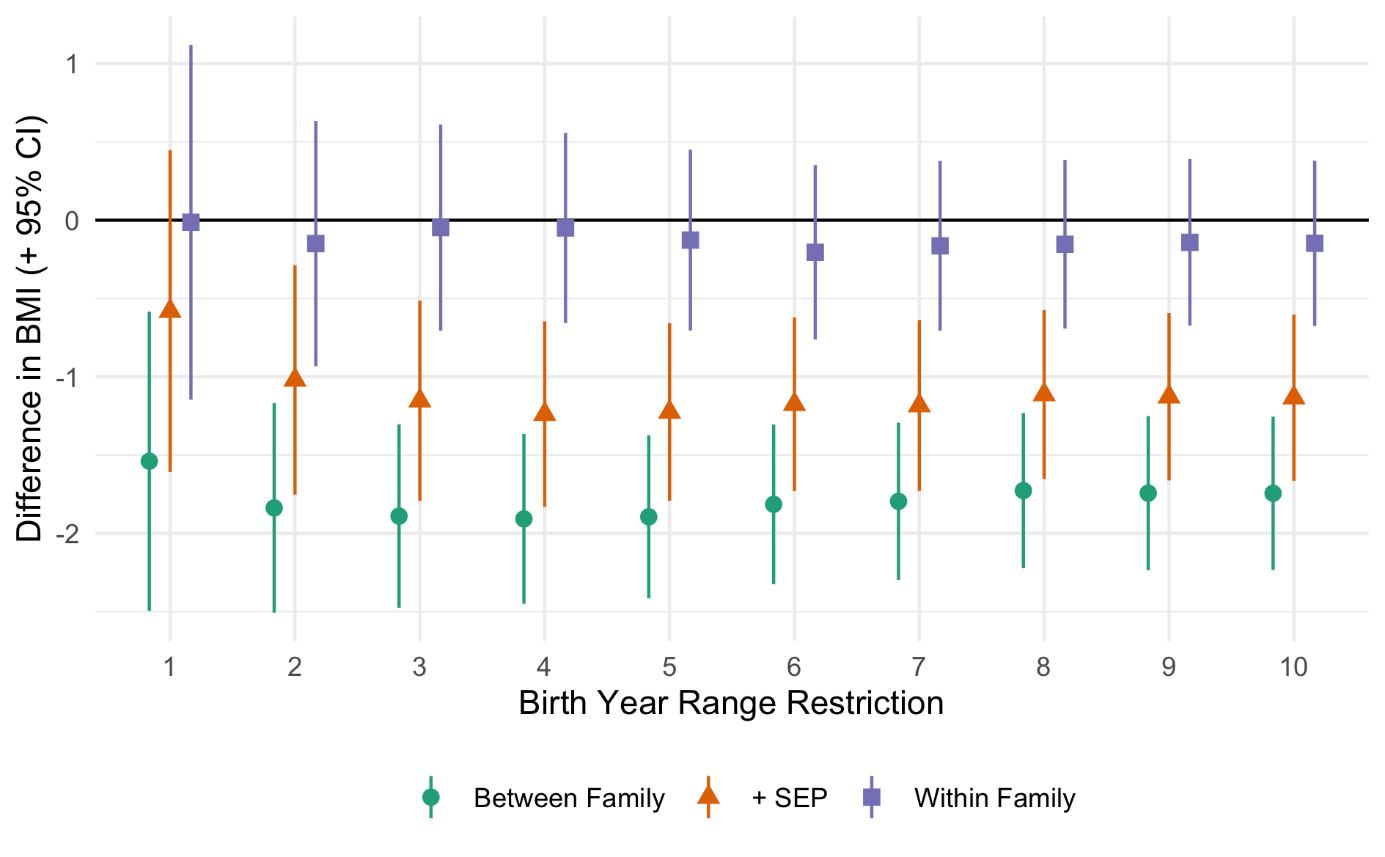

Figure S5: Between and within family associations between cognitive ability (centile rank) and BMI by birth year range used to extract sibling sets. Derived from linear mixed effects models with random intercepts at household and individual level and age (two natural cubic splines), sex, cohort, birth order, ethnic group, and maternal age included as control variables.

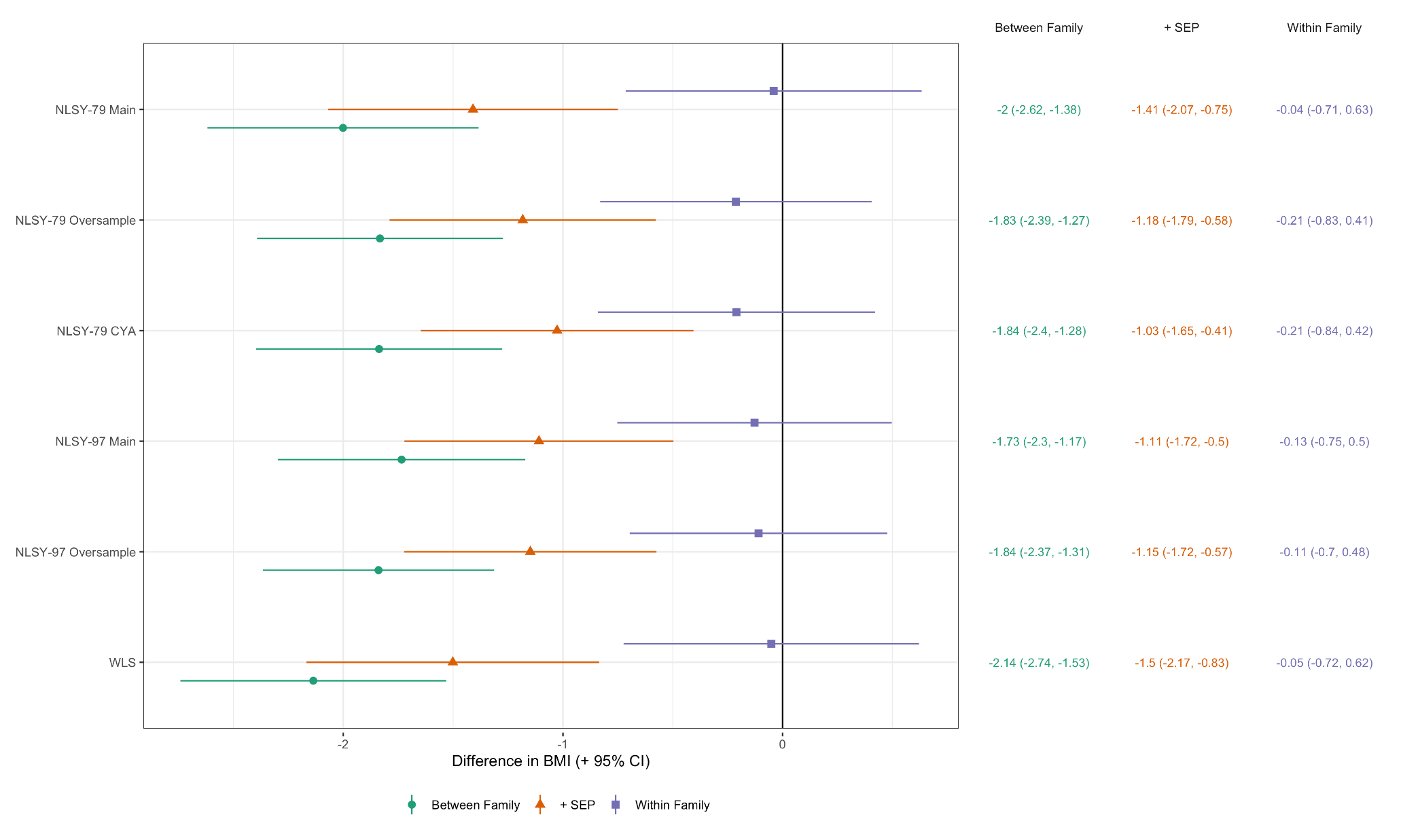

Figure S6: Between and within family associations between cognitive ability (centile rank) and BMI (+ 95% CIs). Models excluding the named cohort (y-axis). Estimates show predicted difference in BMI (kg/m2) comparing individual at the lowest to highest cognition (0th to 100th percentile). Derived from linear mixed effects models with random intercepts at household and individual level and age (two natural cubic splines), sex, cohort, birth order, and maternal age included as control variables.

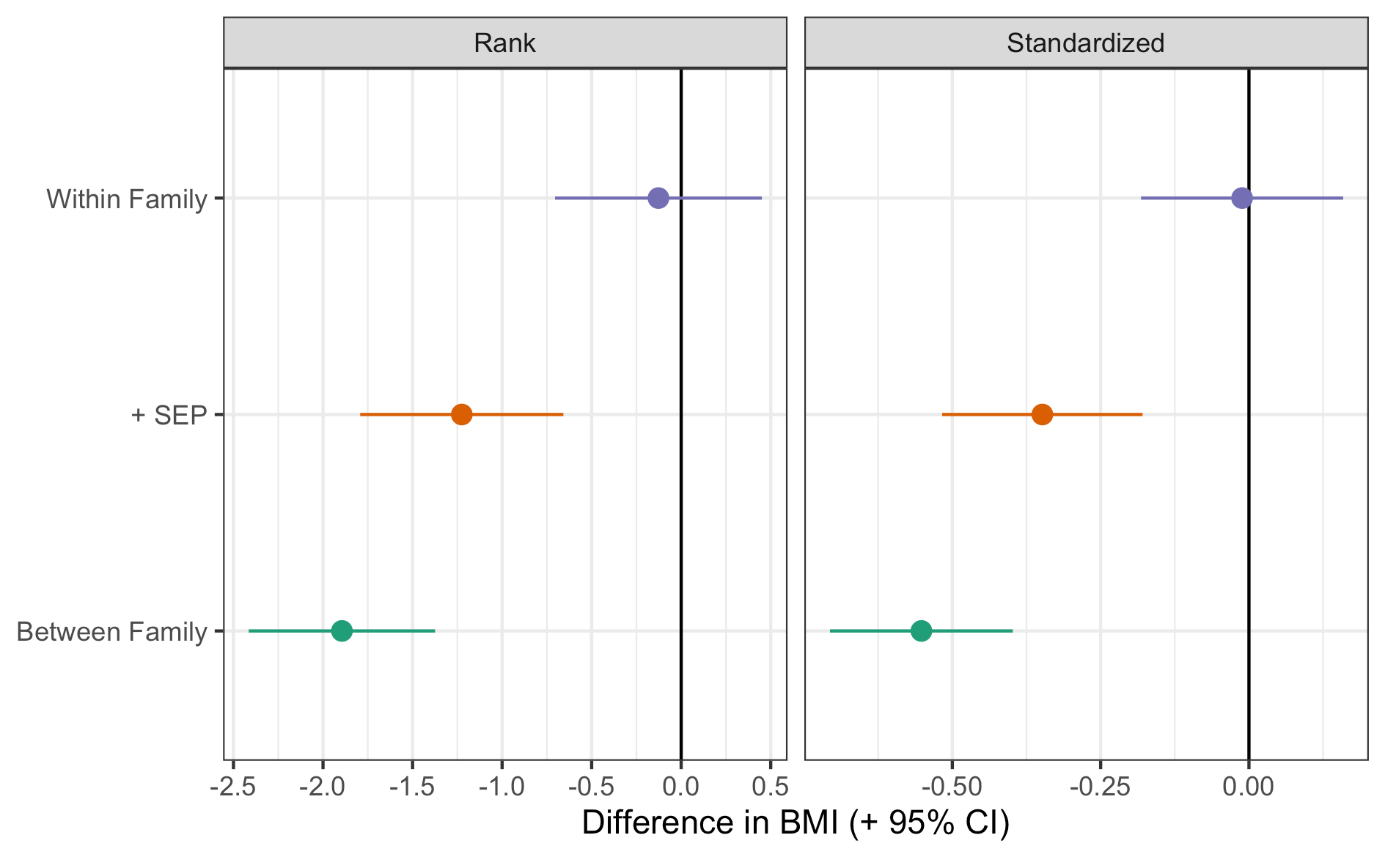

Figure S7: Between and within family associations between cognitive ability and BMI by measure of cognitive ability (centile rank or standardized z-score). Derived from linear mixed effects models with random intercepts at household and individual level and age (two natural cubic splines), sex, cohort, birth order, ethnic group, and maternal age included as control variables.

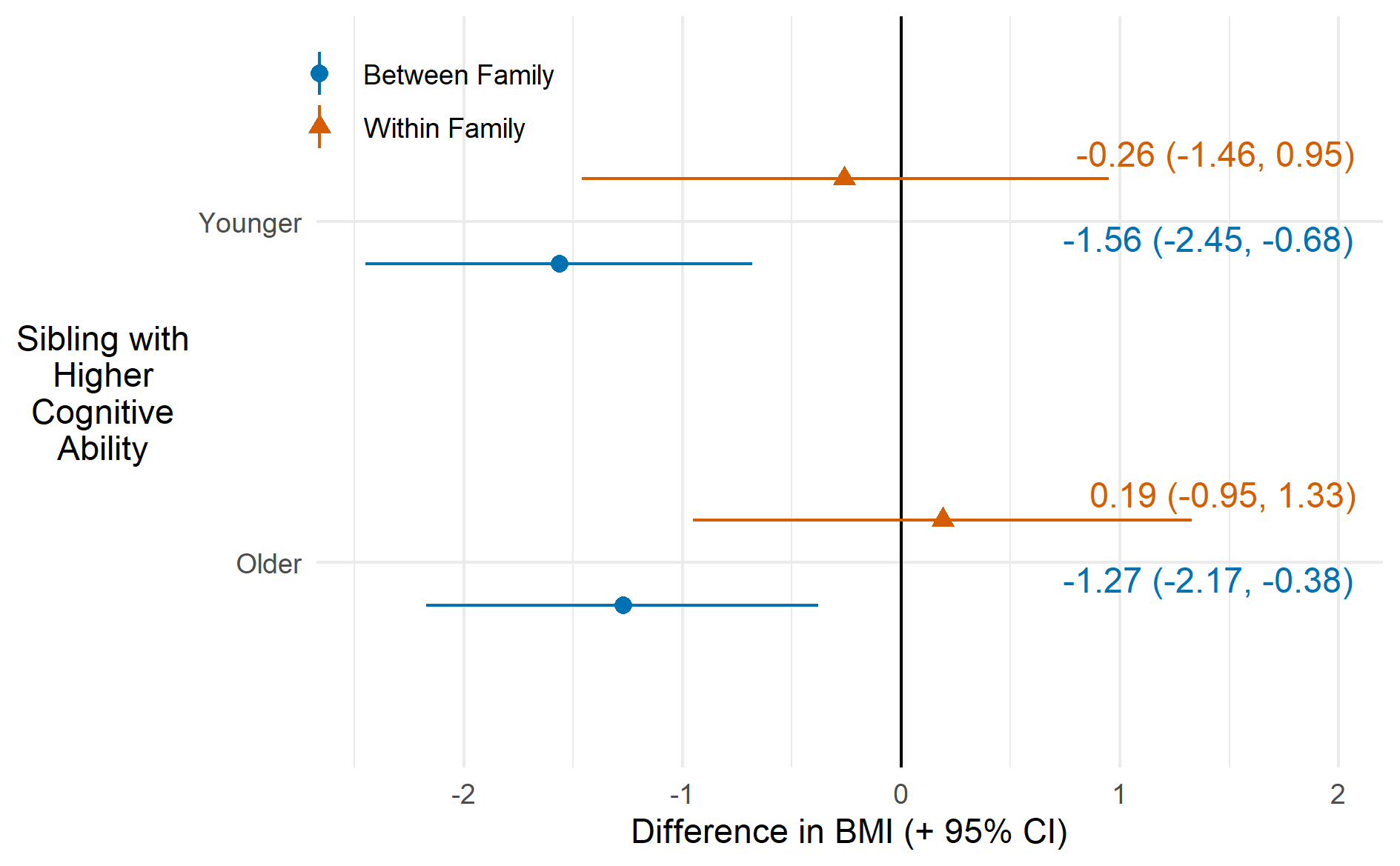

Figure S8: Between and within family associations between cognitive ability and BMI in 2 sibling families, according to whether younger or older sibling had higher cognitive ability score. Derived from linear mixed effects models with random intercepts at household and individual level and age (two natural cubic splines), sex, cohort, birth order, ethnic group, and maternal age included as control variables

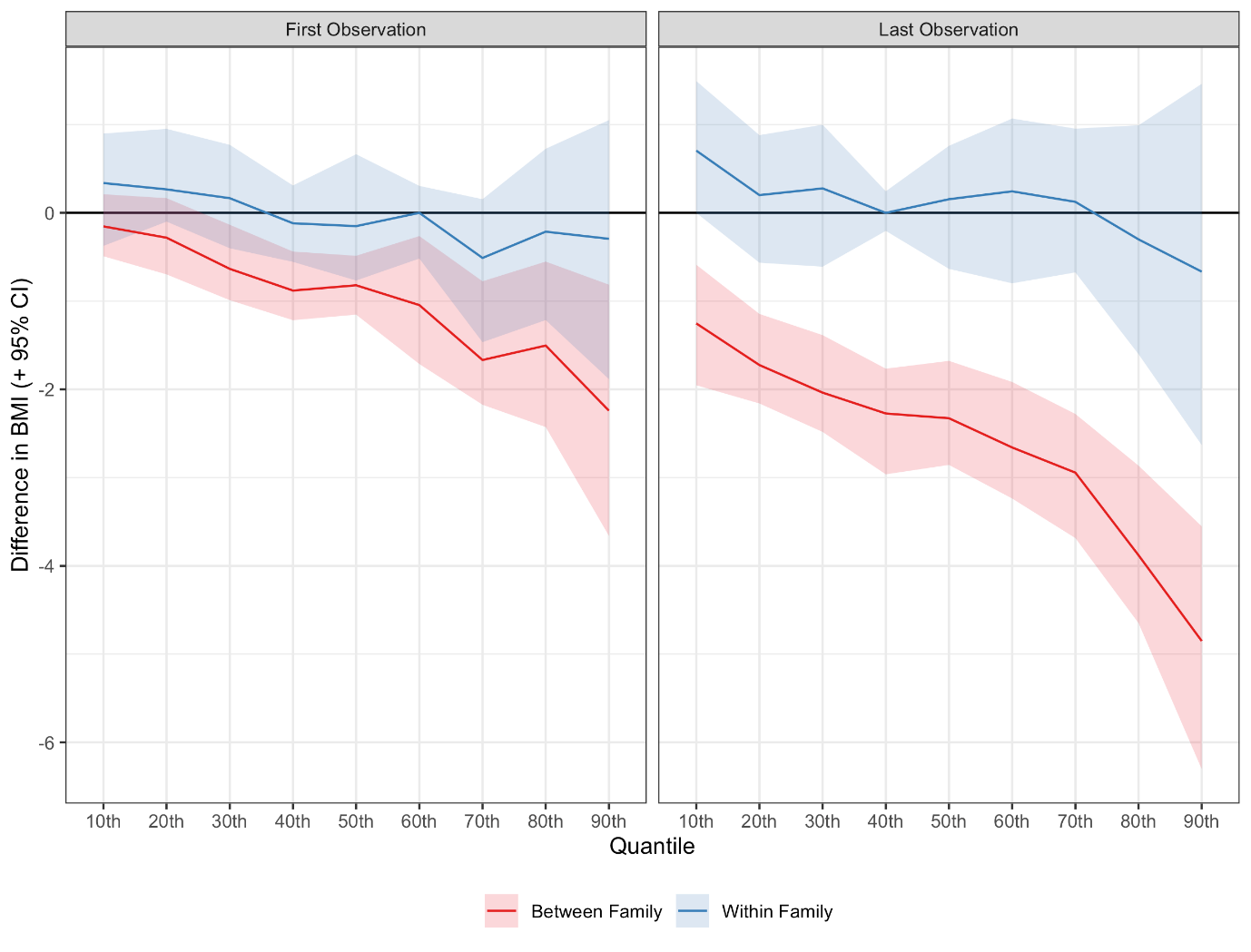

Figure S9: Associations between cognitive ability (percentile) and BMI by quantile of BMI. Derived from residualized quantile regressions using the first (left panel) or last observation (right panel) per individual. Between family effect estimated using one randomly individual per household. Age, maternal age, birth order, sex, SEP, ethnic group and cohort included as control variables in the first stage regressions with household fixed effects also included for within family models. Confidence intervals calculated using cluster-robust bootstrapping (percentile method, 500 replications).

### Tables

Table S1: Sample Selection Flow Table

|  | | Total | NLSY-79 Main | NLSY-79 Oversample | NLSY-79 CYA | NLSY-97 Main | NLSY-97 Oversample | WLS |
| --- | --- | --- | --- | --- | --- | --- | --- | --- |
| All Participants | Households | 29,515 (100%) | 4,012 (100%) | 3,478 (100%) | 4,941 (100%) | 5,184 (100%) | 1,630 (100%) | 10,270 (100%) |
|  | Individuals | 50,981 (100%) | 6,111 (100%) | 5,295 (100%) | 11,545 (100%) | 6,748 (100%) | 2,236 (100%) | 19,046 (100%) |
| Observed Sibling | Households | 14,257 (48.3%) | 1,279 (31.9%) | 857 (24.6%) | 3,068 (62.1%) | 1,249 (24.1%) | 424 (26%) | 7,380 (71.9%) |
|  | Individuals | 31,436 (61.7%) | 3,030 (49.6%) | 2,089 (39.5%) | 7,969 (69%) | 2,639 (39.1%) | 924 (41.3%) | 14,785 (77.6%) |
| Sibling Within +/- 5 Years | Households | 9,782 (68.6%) | 1,262 (98.7%) | 845 (98.6%) | 1,859 (60.6%) | 1,249 (100%) | 424 (100%) | 4,143 (56.1%) |
|  | Individuals | 21,013 (66.8%) | 2,936 (96.9%) | 1,998 (95.6%) | 4,219 (52.9%) | 2,639 (100%) | 924 (100%) | 8,297 (56.1%) |
| Aged 20+ by Final Follow-Up | Households | 9,726 (99.4%) | 1,262 (100%) | 845 (100%) | 1,803 (97%) | 1,249 (100%) | 424 (100%) | 4,143 (100%) |
|  | Individuals | 20,889 (99.4%) | 2,936 (100%) | 1,998 (100%) | 4,095 (97.1%) | 2,639 (100%) | 924 (100%) | 8,297 (100%) |
| Observed BMI @ Age 20+ | Households | 6,665 (68.5%; 68.5%) | 1,245 (98.7%; 98.7%) | 830 (98.2%; 98.2%) | 1,321 (73.3%; 73.3%) | 1,141 (91.4%; 91.4%) | 408 (96.2%; 96.2%) | 1,720 (41.5%; 41.5%) |
|  | Individuals | 14,560 (69.7%; 69.7%) | 2,888 (98.4%; 98.4%) | 1,954 (97.8%; 97.8%) | 2,978 (72.7%; 72.7%) | 2,409 (91.3%; 91.3%) | 887 (96%; 96%) | 3,444 (41.5%; 41.5%) |
| Observed Cognitive Ability | Households | 6,011 (90.2%; 61.8%) | 1,184 (95.1%; 93.8%) | 790 (95.2%; 93.5%) | 1,251 (94.7%; 69.4%) | 943 (82.6%; 75.5%) | 312 (76.5%; 73.6%) | 1,531 (89%; 37%) |
|  | Individuals | 13,146 (90.3%; 62.9%) | 2,737 (94.8%; 93.2%) | 1,859 (95.1%; 93%) | 2,819 (94.7%; 68.8%) | 1,990 (82.6%; 75.4%) | 675 (76.1%; 73.1%) | 3,066 (89%; 37%) |
| Observed Covariates | Households | 5,687 (94.6%; 58.5%) | 1,118 (94.4%; 88.6%) | 751 (95.1%; 88.9%) | 1,251 (100%; 69.4%) | 895 (94.9%; 71.7%) | 299 (95.8%; 70.5%) | 1,373 (89.7%; 33.1%) |
|  | Individuals | 12,427 (94.5%; 59.5%) | 2,569 (93.9%; 87.5%) | 1,760 (94.7%; 88.1%) | 2,819 (100%; 68.8%) | 1,885 (94.7%; 71.4%) | 645 (95.6%; 69.8%) | 2,749 (89.7%; 33.1%) |
| Observed SEP | Households | 5,668 (99.7%; 58.3%) | 1,114 (99.6%; 88.3%) | 750 (99.9%; 88.8%) | 1,251 (100%; 69.4%) | 889 (99.3%; 71.2%) | 291 (97.3%; 68.6%) | 1,373 (100%; 33.1%) |
|  | Individuals | 12,383 (99.6%; 59.3%) | 2,559 (99.6%; 87.2%) | 1,755 (99.7%; 87.8%) | 2,819 (100%; 68.8%) | 1,873 (99.4%; 71%) | 628 (97.4%; 68%) | 2,749 (100%; 33.1%) |
| Discordant Cognitive Ability (Final Sample) | Households | 5,602 (98.8%; 57.6%) | 1,113 (99.9%; 88.2%) | 750 (100%; 88.8%) | 1,246 (99.6%; 69.1%) | 889 (100%; 71.2%) | 291 (100%; 68.6%) | 1,313 (95.6%; 31.7%) |
|  | Individuals | 12,250 (98.9%; 58.6%) | 2,556 (99.9%; 87.1%) | 1,755 (100%; 87.8%) | 2,809 (99.6%; 68.6%) | 1,873 (100%; 71%) | 628 (100%; 68%) | 2,629 (95.6%; 31.7%) |
|  | Observations | 118,355 | 45,842 | 27,332 | 13,954 | 21,332 | 7,266 | 2,629 |

Table S2: Follow-Ups by Cohorts and (Within-Family) Cognitive Ability Group

|  | | | | Follow-Ups | | | | |
| --- | --- | --- | --- | --- | --- | --- | --- | --- |
| Cohort | Group, Cognitive Ability | N | Observations | Mean | SD | Min. | Max. | % Observed |
| NLSY-79 Main | All | 2,556 | 45,842 | 17.94 | 4.73 | 1 | 22 | 85.3% (22.1) |
|  | Below Average | 1,276 | 22,886 | 17.94 | 4.63 | 1 | 22 | 85.3% (21.7) |
|  | Above Average | 1,280 | 22,956 | 17.93 | 4.83 | 1 | 22 | 85.3% (22.6) |
| NLSY-79 Oversample | All | 1,755 | 27,332 | 15.57 | 6.33 | 1 | 22 | 74.1% (29.8) |
|  | Below Average | 897 | 14,041 | 15.65 | 6.32 | 1 | 22 | 74.3% (29.7) |
|  | Above Average | 858 | 13,291 | 15.49 | 6.33 | 1 | 22 | 74% (30) |
| NLSY-79 CYA | All | 2,809 | 13,954 | 4.97 | 1.9 | 1 | 10 | 75.3% (21.7) |
|  | Below Average | 1,396 | 6,894 | 4.94 | 1.9 | 1 | 10 | 75.4% (21.6) |
|  | Above Average | 1,413 | 7,060 | 5 | 1.9 | 1 | 10 | 75.1% (21.8) |
| NLSY-97 Main | All | 1,873 | 21,332 | 11.39 | 3.12 | 1 | 14 | 85.8% (22.6) |
|  | Below Average | 936 | 10,507 | 11.23 | 3.11 | 1 | 14 | 85.6% (22.9) |
|  | Above Average | 937 | 10,825 | 11.55 | 3.12 | 1 | 14 | 86.1% (22.4) |
| NLSY-97 Oversample | All | 628 | 7,266 | 11.57 | 2.86 | 1 | 14 | 86.7% (20.4) |
|  | Below Average | 319 | 3,668 | 11.5 | 2.76 | 1 | 14 | 86.4% (19.9) |
|  | Above Average | 309 | 3,598 | 11.64 | 2.96 | 1 | 14 | 87% (20.9) |
| WLS | All | 2,629 | 2,629 | 1 | 0 | 1 | 1 | 100% (0) |
|  | Below Average | 1,314 | 1,314 | 1 | 0 | 1 | 1 | 100% (0) |
|  | Above Average | 1,315 | 1,315 | 1 | 0 | 1 | 1 | 100% (0) |

Table S3: Descriptive Statistics, Time-Varying Variables

| Cohort | Variable | Group, Cognitive Ability | Mean | Total SD | Between SD | Within SD | Min. | Max. |
| --- | --- | --- | --- | --- | --- | --- | --- | --- |
| NLSY-79 Main | Age | All | 36.7 | 10.8 | 4.0 | 10.4 | 20 | 62 |
|  |  | Below Average | 36.7 | 10.8 | 4.0 | 10.4 | 20 | 62 |
|  |  | Above Average | 36.7 | 10.8 | 4.1 | 10.4 | 20 | 62 |
|  | BMI | All | 26.5 | 5.6 | 4.7 | 3.0 | 13.2 | 69.5 |
|  |  | Below Average | 26.6 | 5.7 | 4.9 | 3.1 | 13.4 | 68.7 |
|  |  | Above Average | 26.4 | 5.5 | 4.6 | 3.0 | 13.2 | 69.5 |
|  | Year | All | 1998.1 | 10.6 | 3.9 | 10.2 | 1981 | 2018 |
|  |  | Below Average | 1998.1 | 10.6 | 3.9 | 10.2 | 1981 | 2018 |
|  |  | Above Average | 1998.1 | 10.6 | 4.0 | 10.2 | 1981 | 2018 |
| NLSY-79 Oversample | Age | All | 36.1 | 10.9 | 5.7 | 10.2 | 20 | 61 |
|  |  | Below Average | 36.2 | 11.0 | 5.7 | 10.2 | 20 | 61 |
|  |  | Above Average | 36.0 | 10.9 | 5.7 | 10.1 | 20 | 61 |
|  | BMI | All | 27.4 | 6.1 | 5.1 | 3.3 | 13.2 | 68.7 |
|  |  | Below Average | 27.3 | 6.1 | 5.1 | 3.3 | 13.2 | 68.7 |
|  |  | Above Average | 27.5 | 6.1 | 5.2 | 3.3 | 14.8 | 68.7 |
|  | Year | All | 1997.5 | 10.7 | 5.5 | 10.0 | 1981 | 2018 |
|  |  | Below Average | 1997.5 | 10.8 | 5.5 | 10.1 | 1981 | 2018 |
|  |  | Above Average | 1997.5 | 10.7 | 5.5 | 10.0 | 1981 | 2018 |
| NLSY-79 CYA | Age | All | 26.4 | 4.9 | 2.5 | 4.4 | 20 | 46 |
|  |  | Below Average | 26.3 | 4.9 | 2.4 | 4.3 | 20 | 46 |
|  |  | Above Average | 26.4 | 4.9 | 2.5 | 4.4 | 20 | 46 |
|  | BMI | All | 27.2 | 6.3 | 5.6 | 2.6 | 13.6 | 66.4 |
|  |  | Below Average | 27.2 | 6.3 | 5.7 | 2.6 | 13.6 | 63 |
|  |  | Above Average | 27.2 | 6.2 | 5.6 | 2.7 | 14.4 | 66.4 |
|  | Year | All | 2010.3 | 5.2 | 3.6 | 4.2 | 1994 | 2018 |
|  |  | Below Average | 2010.3 | 5.2 | 3.6 | 4.1 | 1994 | 2018 |
|  |  | Above Average | 2010.2 | 5.2 | 3.6 | 4.2 | 1994 | 2018 |
| NLSY-97 Main | Age | All | 27.3 | 5.0 | 1.8 | 4.8 | 20 | 40 |
|  |  | Below Average | 27.2 | 5.0 | 1.8 | 4.8 | 20 | 40 |
|  |  | Above Average | 27.4 | 5.0 | 1.8 | 4.8 | 20 | 40 |
|  | BMI | All | 27.1 | 6.3 | 5.8 | 2.6 | 13.2 | 68.6 |
|  |  | Below Average | 27.2 | 6.3 | 5.7 | 2.6 | 13.2 | 68.6 |
|  |  | Above Average | 27.0 | 6.4 | 5.9 | 2.6 | 14.6 | 66.9 |
|  | Year | All | 2009.4 | 4.9 | 1.6 | 4.8 | 2002 | 2019 |
|  |  | Below Average | 2009.5 | 4.9 | 1.6 | 4.8 | 2002 | 2019 |
|  |  | Above Average | 2009.3 | 4.9 | 1.6 | 4.8 | 2002 | 2019 |
| NLSY-97 Oversample | Age | All | 27.5 | 5.0 | 1.6 | 4.8 | 20 | 40 |
|  |  | Below Average | 27.5 | 5.0 | 1.5 | 4.8 | 20 | 40 |
|  |  | Above Average | 27.5 | 5.0 | 1.6 | 4.9 | 20 | 40 |
|  | BMI | All | 28.7 | 6.6 | 5.9 | 2.9 | 14.1 | 64.8 |
|  |  | Below Average | 28.8 | 6.7 | 6.1 | 3.0 | 14.1 | 64.8 |
|  |  | Above Average | 28.5 | 6.5 | 5.7 | 2.9 | 14.1 | 61.3 |
|  | Year | All | 2009.4 | 4.9 | 1.4 | 4.8 | 2002 | 2019 |
|  |  | Below Average | 2009.5 | 4.9 | 1.3 | 4.8 | 2002 | 2019 |
|  |  | Above Average | 2009.4 | 4.9 | 1.5 | 4.8 | 2002 | 2019 |
| WLS | Age | All | 54.2 | 2.4 | 2.4 | 0.0 | 49 | 62.1 |
|  |  | Below Average | 54.2 | 2.4 | 2.4 | 0.0 | 49 | 61.2 |
|  |  | Above Average | 54.2 | 2.4 | 2.4 | 0.0 | 49.1 | 62.1 |
|  | BMI | All | 26.6 | 4.4 | 4.4 | 0.0 | 16.6 | 50.5 |
|  |  | Below Average | 26.7 | 4.5 | 4.5 | 0.0 | 16.6 | 50.5 |
|  |  | Above Average | 26.5 | 4.4 | 4.4 | 0.0 | 18 | 46.8 |
|  | Year | All | 1993.3 | 0.8 | 0.8 | 0.0 | 1992 | 1994 |
|  |  | Below Average | 1993.3 | 0.8 | 0.8 | 0.0 | 1992 | 1994 |
|  |  | Above Average | 1993.3 | 0.8 | 0.8 | 0.0 | 1992 | 1994 |

Table S4: Residualized Quantile Regression Results

| Quantile | Within Family | Between Family | excl. NLSY-79 Main | excl. NLSY-79 Oversample | excl. NLSY-79 CYA | excl. NLSY-97 Main | excl. NLSY-97 Oversample | excl. WLS |
| --- | --- | --- | --- | --- | --- | --- | --- | --- |
| 10th | 0.34 (-0.37, 0.9) | -0.15 (-0.49, 0.21) | 0.27 (-0.34, 1) | 0.11 (-0.38, 0.99) | 0.13 (-0.44, 0.87) | 0.32 (-0.39, 0.94) | 0.31 (-0.32, 0.91) | 0.37 (-0.5, 1.12) |
| 20th | 0.27 (-0.1, 0.95) | -0.28 (-0.7, 0.17) | 0.57 (-0.3, 1.14) | 0.36 (-0.16, 0.94) | 0.38 (-0.08, 1.07) | 0.2 (-0.28, 0.96) | 0.33 (-0.03, 0.95) | 0.25 (-0.44, 0.76) |
| 30th | 0.17 (-0.4, 0.77) | -0.63 (-0.99, -0.14) | -0.11 (-0.66, 0.57) | 0.03 (-0.48, 0.6) | 0.12 (-0.52, 0.95) | 0 (-0.59, 0.7) | 0.08 (-0.33, 0.81) | 0.27 (-0.11, 0.74) |
| 40th | -0.12 (-0.56, 0.31) | -0.88 (-1.22, -0.44) | 0 (-0.8, 0.65) | -0.04 (-0.67, 0.52) | -0.14 (-0.63, 0.45) | -0.09 (-0.71, 0.37) | -0.23 (-0.68, 0.38) | 0 (-0.49, 0.57) |
| 50th | -0.15 (-0.77, 0.66) | -0.82 (-1.15, -0.49) | -0.04 (-0.42, 0.33) | 0 (-0.74, 0.58) | -0.09 (-0.84, 0.57) | -0.1 (-0.79, 0.66) | -0.05 (-0.63, 0.57) | 0 (-0.76, 0.74) |
| 60th | 0 (-0.52, 0.31) | -1.05 (-1.71, -0.26) | 0 (-0.97, 0.73) | 0 (-1.16, 0.74) | -0.04 (-0.81, 0.68) | -0.05 (-0.58, 0.51) | 0 (-0.63, 0.54) | -0.04 (-0.54, 0.3) |
| 70th | -0.51 (-1.46, 0.16) | -1.67 (-2.17, -0.78) | -0.62 (-1.52, 0.7) | -0.16 (-1.57, 0.08) | -1.03 (-1.57, 0.57) | -0.62 (-1.6, 0.6) | -0.72 (-1.59, 0.37) | -0.36 (-1.39, 0.49) |
| 80th | -0.21 (-1.21, 0.73) | -1.5 (-2.43, -0.55) | -0.11 (-1.36, 0.95) | -0.48 (-1.49, 0.75) | -0.32 (-1.61, 0.8) | -0.43 (-1.48, 0.68) | -0.27 (-1.29, 0.78) | -0.1 (-1.26, 0.52) |
| 90th | -0.29 (-1.88, 1.05) | -2.24 (-3.66, -0.81) | -0.84 (-3.03, 0.89) | -0.56 (-2.27, 1.2) | -0.03 (-1.66, 1.59) | -0.99 (-2.35, 0.37) | -0.36 (-1.83, 1.08) | 0 (-2.38, 2.12) |

### References

Altschul, D. M., Wraw, C., Gale, C. R., & Deary, I. J. (2019). How youth cognitive and sociodemographic factors relate to the development of overweight and obesity in the UK and the USA: A prospective cross-cohort study of the National Child Development Study and National Longitudinal Study of Youth 1979. *BMJ Open*, *9*(12), e033011. https://doi.org/10.1136/bmjopen-2019-033011

Herd, P., Carr, D., & Roan, C. (2014). Cohort Profile: Wisconsin longitudinal study (WLS). *International Journal of Epidemiology*, *43*(1), 34–41. https://doi.org/10.1093/ije/dys194

Herrnstein, R. J., & Murray, C. A. (1996). *The bell curve: Intelligence and class structure in American life* (1st Free Press pbk. ed). Simon & Schuster.

Michael, R. T., & Pergamit, M. R. (2001). The National Longitudinal Survey of Youth, 1997 Cohort. *The Journal of Human Resources*, *36*(4), 628. https://doi.org/10.2307/3069636

Rasmusen, E. (2007, February 5). *The Bell Curve Page*. http://www.rasmusen.org/xpacioli/bellcurve/bellcurve.htm

Rodgers, J. L., Beasley, W. H., Bard, D. E., Meredith, K. M., Hunter, M. D., Johnson, A. B., Buster, M., Li, C., May, K. O., Garrison, S. M., Miller, W. B., van den Oord, E., & Rowe, D. C. (2016). The NLSY Kinship Links: Using the NLSY79 and NLSY-Children Data to Conduct Genetically-Informed and Family-Oriented Research. *Behavior Genetics*, *46*(4), 538–551. https://doi.org/10.1007/s10519-016-9785-3

Rohrer, J. M., Egloff, B., & Schmukle, S. C. (2015). Examining the effects of birth order on personality. *Proceedings of the National Academy of Sciences*, *112*(46), 14224–14229. https://doi.org/10.1073/pnas.1506451112

Rothstein, D. S., Carr, D., & Cooksey, E. (2019). Cohort Profile: The National Longitudinal Survey of Youth 1979 (NLSY79). *International Journal of Epidemiology*, *48*(1), 22–22e. https://doi.org/10.1093/ije/dyy133

Wraw, C., Deary, I. J., Der, G., & Gale, C. R. (2018). Maternal and offspring intelligence in relation to BMI across childhood and adolescence. *International Journal of Obesity*, *42*(9), 1610–1620. https://doi.org/10.1038/s41366-018-0009-1

Wraw, C., Deary, I. J., Gale, C. R., & Der, G. (2015). Intelligence in youth and health at age 50. *Intelligence*, *53*, 23–32. https://doi.org/10.1016/j.intell.2015.08.001
